# Clinically Generalisable End-to-End Graph Learning for CT Image-Based Multitask Stroke Diagnosis

**DOI:** 10.64898/2026.08.26.26360026

**Authors:** Zhicheng Lu, Shahadat Uddin, Sergio Uribe, Sam White, Rodrigo Tomazini Martins, Shayne Chau, Abu Syed Md. Mosaddek, Md. Siddiqul Islam, Nusratun Nahar, AKM Azad, K. M. Nazmul Hossain, Habib Sadat Choudhury, K.M. Rakibul Hasan, Nabil Mosaddek, Samia Rahman, Md. Mostaque Hossain, K.M. Mehedi Hasan Sizar, Claudio Angione, Pietro Liò, Md Tauhidul Islam, Mohammad Ali Moni

**Affiliations:** Rural Health Research Institute, Charles Sturt University, Orange, NSW 2800, Australia; School of Electrical and Computer Engineering, Faculty of Engineering, The University of Sydney, Darlington, NSW 2006, Australia; Department of Medical Imaging and Radiation Science, School of Primary and Allied health Care, Faculty of Medicine Nursing and Health Science, Monash University, Victoria 3800, Australia; SA Medical Imaging, South Australia, Australia; Faculty of Science and Health, School of Dentistry and Medical Sciences, Charles Sturt University, Wagga Wagga, NSW 2650, Australia; Mater Misericordiae Hospital, Queensland, Australia; Wagga Wagga Base Hospital, Wagga Wagga, NSW 2650, Australia; Murrumbidgee Local Health District, Wagga Wagga, NSW 2650, Australia; Department of Pharmacology, Uttara Adhunik Medical College, Dhaka, Bangladesh; Quest Bangladesh Dhaka, Bangladesh; Department of Pharmacy, American International University-Bangladesh, Dhaka, Bangladesh; Department of Pharmacy, Southeast University, Dhaka, Bangladesh; Department of Mathematics and Statistics, Faculty of Science, Imam Mohammad Ibn Saud Islamic University (IMSIU), Riyadh 13318, Saudi Arabia; University Medical Center, Jahangirnagar University, Dhaka, Bangladesh; Department of Biochemistry, International Medical College, Gazipur, Bangladesh; Paediatric Neuro Care & Research (PNR), Dhaka, Bangladesh; Department of Biomedical Engineering, Military Institute of Science and Technology (MIST), Dhaka, Bangladesh; Popular Diagnostic Centre, Dhaka, Bangladesh; Department of Radiology, International Medical College, Gazipur, Bangladesh; IbnaSina Diagnostic & Imaging Center, Dhaka, Bangladesh; School of Computing, Engineering & Digital Technologies, Teesside University, UK; Department of Computer Science and Technology, University of Cambridge, Cambridge, UKxg112; Department of Radiation Oncology, Stanford University, Stanford, CA, USA; Artificial Intelligence and Cyber Futures Institute, Charles Sturt University, Bathurst, NSW 2795, Australia; School of Health and Rehabilitation Sciences, The University of Queensland, St Lucia, Brisbane, QLD 4072, Australia

## Abstract

Stroke remains a leading cause of mortality and long-term disability worldwide, yet rapid diagnosis is often limited by the shortage of trained radiologists, particularly in resource-constrained settings. Automated analysis of CT imaging offers a potential solution, but existing methods often struggle to achieve clinically generalisable performance while jointly addressing multiple diagnostic tasks. Here we present the Intelligent Integrated Stroke Diagnosis System (IISDS), an end-to-end deep learning framework built upon StrokeGNN, a graph-based architecture that integrates 3D contextual feature extraction with U-Net-based 2D lesion segmentation to enable comprehensive stroke analysis from non-contrast CT scans. IISDS performs stroke subtype classification, lesion segmentation and lesion volume estimation within a unified pipeline. To develop and validate the system, we collected and curated BGD-ISD through a collaboration between AI researchers, neurologists, radiologists and clinicians, resulting in a large multi-centre dataset comprising 1,507 CT scans from 597 stroke cases acquired across six hospitals and medical centres in Bangladesh. Across BGD-ISD and multiple publicly available datasets, IISDS achieves state-of-the-art performance on all tasks, improving segmentation accuracy by ≥ 0.011 Dice score, reducing lesion volume estimation error by ≥ 0.3 average symmetric surface distance (ASSD), and increasing classification performance by ≥ 0.018 area under the receiver operating characteristic curve (AUC) compared with existing approaches. These results demonstrate the potential of graph-based deep learning to enable clinically generalisable, automated and scalable stroke diagnosis from CT imaging, supporting rapid clinical decision-making, particularly in healthcare environments with limited access to expert radiological interpretation. The BGD-ISD dataset will be made openly available to the research community upon publication at: https://github.com/Zhicheng-Lu/stroke_ct.

## Introduction

Stroke is a major global health challenge and remains one of the leading causes of mortality and long-term disability worldwide^1^. Each year, more than 15 million people experience a stroke, with approximately one-third resulting in death and another third leading to permanent physical or cognitive impairment^1^. Several risk factors such as hypertension, diabetes, atrial fibrillation, and aging contribute substantially to the global burden of stroke^2^. Notably, nearly 75% of stroke cases occur in individuals aged 65 years or older^3^. As populations continue to age globally, the incidence and healthcare burden associated with stroke are expected to increase substantially. Computed tomography (CT) is the primary imaging modality used in emergency stroke assessment because of its speed, accessibility and ability to rapidly differentiate hemorrhagic from ischemic stroke^4,5^. However, accurate interpretation of CT scans requires considerable expertise, and subtle imaging findings can be difficult to detect, particularly in early-stage ischemic stroke. This diagnostic bottleneck highlights the urgent need for scalable solutions that can support and extend clinical diagnostic capacity^6^.

Artificial intelligence (AI) methods have emerged as a promising approach to assist clinicians by providing rapid, consistent and comprehensive analysis of medical images. Increasingly, collaborations between AI researchers and clinical institutions enable the development of practical AI systems that support real-world diagnostic workflows^7^. In the context of stroke diagnosis, many studies have focused on distinguishing ischemic and hemorrhagic stroke types because these conditions often present with similar clinical symptoms (such as acute confusion, speech impairment, focal neurological deficits and severe headache), yet require fundamentally different treatment strategies^8–10^. Recent advances in machine learning (ML) and deep learning (DL)^11–14^ have demonstrated performance approaching that of expert radiologists in stroke classification tasks. Beyond classification, lesion segmentation has also received considerable attention, as accurate delineation of stroke lesions enables localization of pathology, estimation of lesion burden and derivation of clinically relevant metrics such as the Alberta Stroke Program Early CT Score (ASPECTS), which is used to guide treatment decisions. Numerous segmentation approaches^15–21^ have achieved strong performance and, in some cases outperform human annotations.

Despite these advances, several challenges remain. Most existing approaches address individual diagnostic tasks in isolation, limiting their utility for comprehensive clinical decision support. Furthermore, the complex three-dimensional spatial structure of CT volumes is often underutilized by conventional architectures. Few existing systems integrate volumetric feature representation with lesion-level analysis while simultaneously supporting multiple clinically relevant tasks. In addition, reliable estimation of stroke severity remains an important component of treatment prioritization. Clinical scales such as the National Institutes of Health Stroke Scale (NIHSS)^22^ are widely used but primarily reflect observable neurological deficits rather than imaging-derived characteristics of the lesion itself. Quantitative imaging biomarkers, such as lesion volume, may therefore provide complementary information for assessing stroke severity.

Here we present an AI-driven Intelligent Integrated Stroke Diagnosis System (IISDS) for automated diagnosis from CT imaging. IISDS performs stroke subtype classification, lesion segmentation and lesion volume estimation within a unified computational framework. At the core of the system is StrokeGNN, a graph-based deep learning architecture that integrates volumetric representation learning using three-dimensional graph neural networks with two-dimensional U-Net–based segmentation features. This hybrid design enables efficient modeling of spatial relationships in CT volumes while preserving detailed lesion boundary information. To support model development and evaluation, our multi-disciplinary team of AI researchers, neurologists, radiologists and clinicians collect and curate BGD-ISD, a multi-centre clinical dataset consisting of 1,507 CT scans from 597 stroke cases collected across six hospitals and medical centres in Bangladesh. The dataset captures diverse real-world clinical conditions and imaging variability, providing a robust benchmark for evaluating automated stroke analysis systems. To facilitate reproducibility and further research, BGD-ISD will be publicly released at the project repository: https://github.com/Zhicheng-Lu/stroke_ct. Experimental results demonstrate that IISDS achieves state-of-the-art performance across stroke classification, lesion segmentation and lesion volume estimation tasks, with performance comparable to expert radiologists. By integrating multiple diagnostic components within a unified graph-based framework, IISDS moves beyond task-specific AI models and provides a scalable approach for automated stroke analysis from CT imaging, with potential to support clinical decision-making in real-world healthcare environments.

## Results

### IISDS-enabled doctor-level lesion segmentation

As a core component of IISDS, stroke segmentation is trained using multiple publicly available datasets and serves as the foundation for downstream volume estimation and classification. For ischemic stroke, AISD^15^ contains 397 CT scans of acute ischemic stroke with a slice thickness of 5 mm. Ischemic lesions were manually annotated by a radiologist based on MRI findings and independently reviewed by a senior radiologist. Among the 397 scans, 345 are used for training and validating, while the remaining 52 are reserved for testing^15^. For hemorrhagic stroke, PhysioNet-ICH^23^ provides 82 CT scans with a slice thickness of 5 mm, including 36 scans from hemorrhagic patients. Among all patients, 46 are male and 36 are female, with a mean age of 2.8 years and a standard deviation of 19.5. The CT scans were reviewed and annotated by two radiologists. BHSD^20^ is a 192-scan subset of the original RSNA dataset^24^, where pixel-level multi-class and multi-rater annotations were performed by four medical imaging professionals and subsequently reviewed by an experienced radiologist. Seg-CQ500^21^ consists of 51 annotated scans from the original CQ500 dataset^25^, including 41 scans with ≤ 0.625 mm slice thickness and 10 scans ranging from 3 mm to 5mm. Annotations were manually generated by one radiologist and verified by an expert neuroradiologist. Details of the datasets are illustrated in Supplementary Table S6.

To ensure consistency across multi-site datasets, all CT scans are resampled to an isotropic in-plane resolution of 0.5×0.5 mm while preserving their original through-plane resolution. This choice was motivated by two factors: (1) The majority of our available datasets were originally acquired at approximately this pixel spacing (512×512 matrix size); (2) A 5 mm slice interval remains the standard protocol in routine clinical stroke imaging^26^, particularly in emergency and resource-limited settings. By retaining each scan’s native through-plane resolution, we avoid artificially upsampling thick-slice data and preserve the intrinsic variability present in real-world clinical acquisitions. Only the in-plane resolution is normalized, which reduces heterogeneity across cohorts while maintaining the z-axis. Although this approach results in thinner-slice datasets, such as Seg-CQ500, that have substantially more slices, it provides IISDS with a different view of high-resolution CT scans. To increase the capability of the model, we cut the original input R^*D*×*W*×*H*^ into smaller R^30×256×256^ patches with a minimal 32 pixels of overlap between any two neighboring patches. This patch-based design improves GPU memory efficiency while preserving sufficient contextual continuity across slices, enabling robust learning of volumetric stroke patterns. For scans that have less than 30 slices, the through-plane resolution keeps unchanged.

Firstly, we evaluate the proposed model using a five-fold cross-validation protocol. To address class imbalance and data deficiency, we augment the training sets of PhysioNet-ICH and Seg-CQ500 by factors of 3 and 2, respectively. We use four of the five folds (∼640 augmented scans from 800) for training and validation, and the remaining fold (∼ 135 non-augmented scans from 677) for testing. Detailed data distribution is illustrated in Figure 2a (see Supplementary Figure S2a for more details), while slice distribution is shown in Figure 2b. For performance evaluation, we employ six quantitative metrics: Dice score (F1), Intersection over Union (IoU), precision, recall, Average Symmetric Surface Distance (ASSD), and Hausdorff distance (HD).

**Figure 1.**
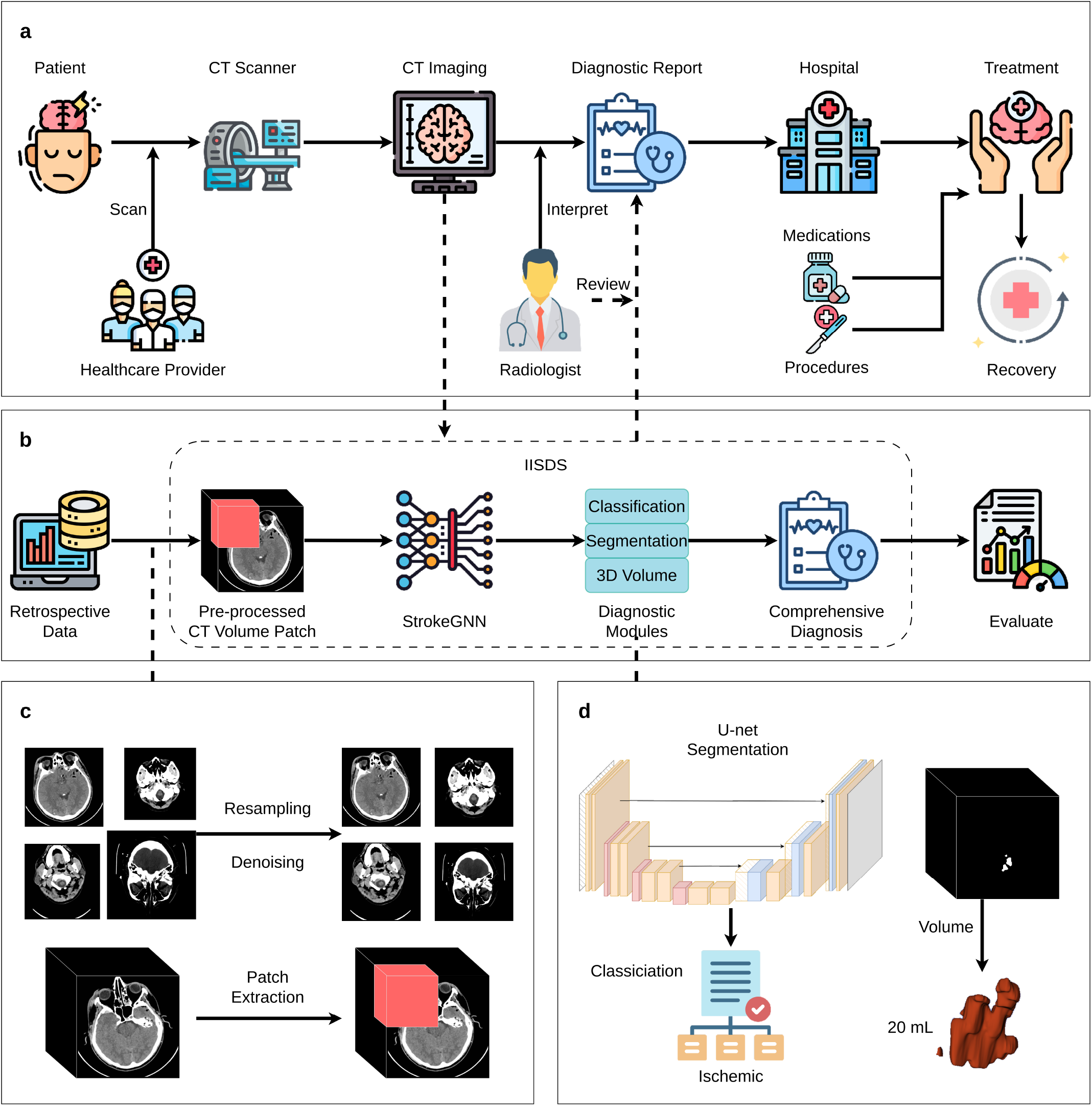
Overview of the proposed workflow. (a) Traditional clinical pathway: suspected stroke patients undergo CT imaging, after which radiologists manually review the scans and generate diagnostic reports to guide treatment decisions. (b) AI-enabled workflow: CT images are automatically processed by the pre-trained IISDS to produce comprehensive diagnostic outputs, which are subsequently reviewed and confirmed by radiologists. (c) Pre-processing pipeline, including intensity normalization, resampling to a unified spatial resolution, and patch-based volume construction. (d) Multi-task diagnostic inference, comprising stroke subtype classification, lesion segmentation, and 3D lesion volume estimation. Detailed model architecture and implementation are provided in Supplementary Figure S1 and Supplementary Table S2, with full methodological descriptions of the pre-processing and multi-task learning framework in Section Methods.

**Figure 2.**
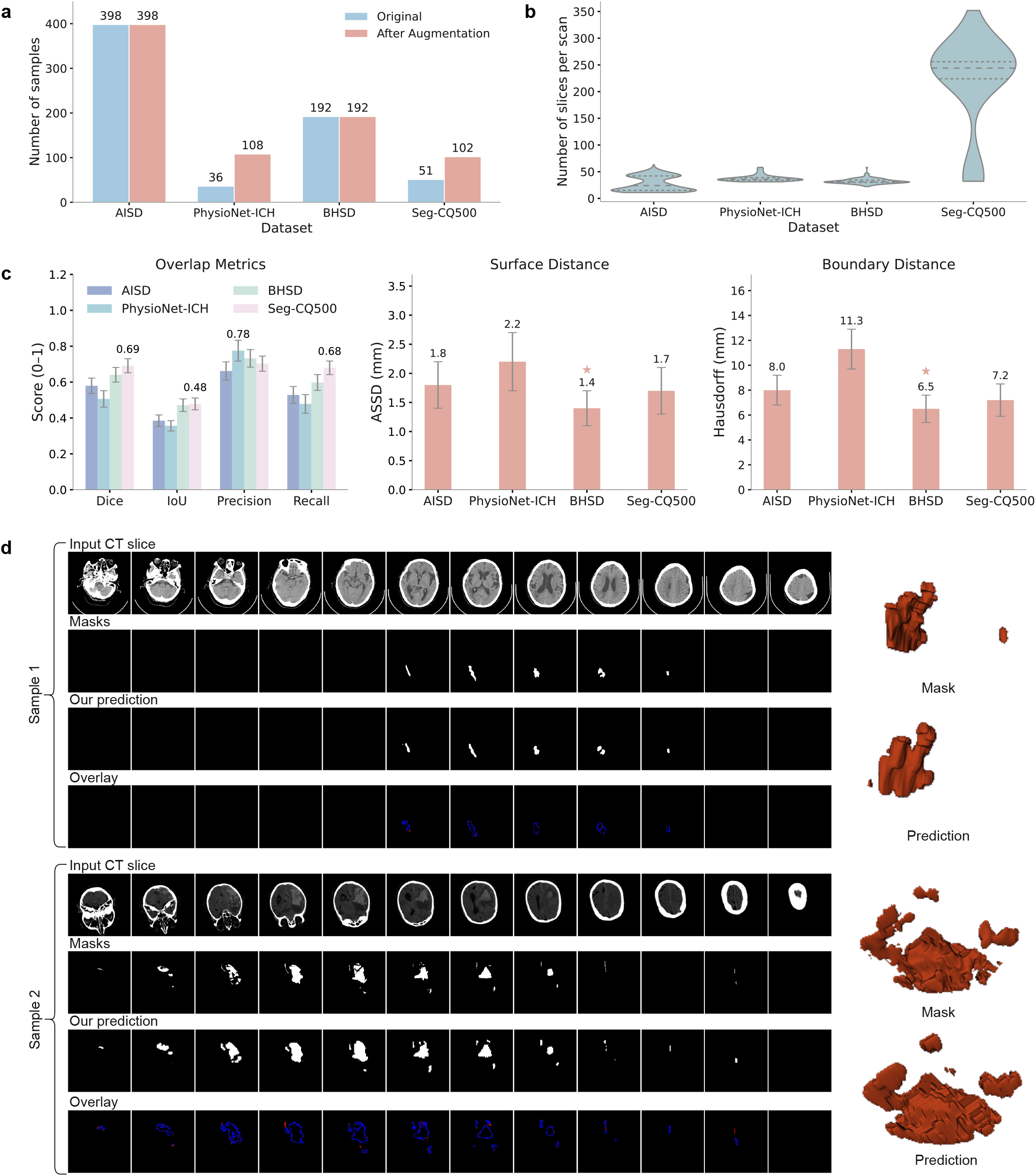
Experimental results for stroke lesion segmentation using a cross-validation setup. (a) Dataset distribution before and after augmentation. (b) Slice count (z-axis) distribution for each dataset. (c) Quantitative testing results including Dice, IoU, precision, recall, ASSD, and Hausdorff distance across the original four datasets. (d) Left: slice-level qualitative results for two representative cases (one AISD, one Seg-CQ500), showing original CT, ground-truth annotation, model prediction, and overlay comparison (top to bottom); right: 3D visualization of lesion volumes for the same cases.

On average, training converges after approximately 202 epochs (or 5900 minutes) across the five folds. Supplementary Figure S2b illustrates the training and validation loss curves for fold 1, where training terminates around the 200^th^ epoch according to the early stop criteria. Quantitative results averaged across the five folds are shown in Figure 2c (see Supplementary Figure S2c for more details), yielding an overall Dice score of 0.6434. Dice scores for individual datasets are 0.5793 (AISD), 0.5060 (PhysioNet-ICH), 0.6406 (BHSD), and 0.6910 (Seg-CQ500), respectively. Notably, the model achieves relatively high precision (0.7053) but lower recall (0.5983), likely because scan-level evaluation penalizes false-negative samples with tiny lesion regions. Qualitative segmentation examples from one ischemic case (AISD) and one hemorrhagic case (Seg-CQ500) are presented in Figure 2d in both 2D and 3D views. These examples confirm that StrokeGNN captures both lesion shape and volume with strong subjective fidelity.

To further validate the proposed model, we benchmark its performance against state-of-the-art segmentation methods on AISD and PhysioNet-ICH datasets. We include the full version of AISD, PhysioNet-ICH, BHSD, and Seg-CQ500. The test sets consist of 52 AISD scans and all 36 PhysioNet-ICH scans according to the setup from Liang *et al*.^15^ and Hssayeni *et al*.^18^. Additionally, we augment and BHSD as before to improve generalizability. Supplementary Figure S3a shows the data distribution. For training, we randomly hold out 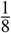 of AISD samples for validation. Training converges after 218 epochs (6419 minutes), with stable validation performance across datasets (Supplementary Figure S3b).

We compare our model with existing methods on AISD, including UNet^27^, SEAN^15^, ADN^16^, and Xu *et al*.^17^. As shown in Figure 3a (see Supplementary Figure S3c for more details), our model achieves the highest Dice score of 0.5889, outperforming U-Net (0.4588), SEAN (0.5784), ADN (0.5245), and Xu *et al*. (0.5866). Our method also achieves the highest IoU, precision, ASSD, and HD, demonstrating its superior lesion-detection capability.

**Figure 3.**
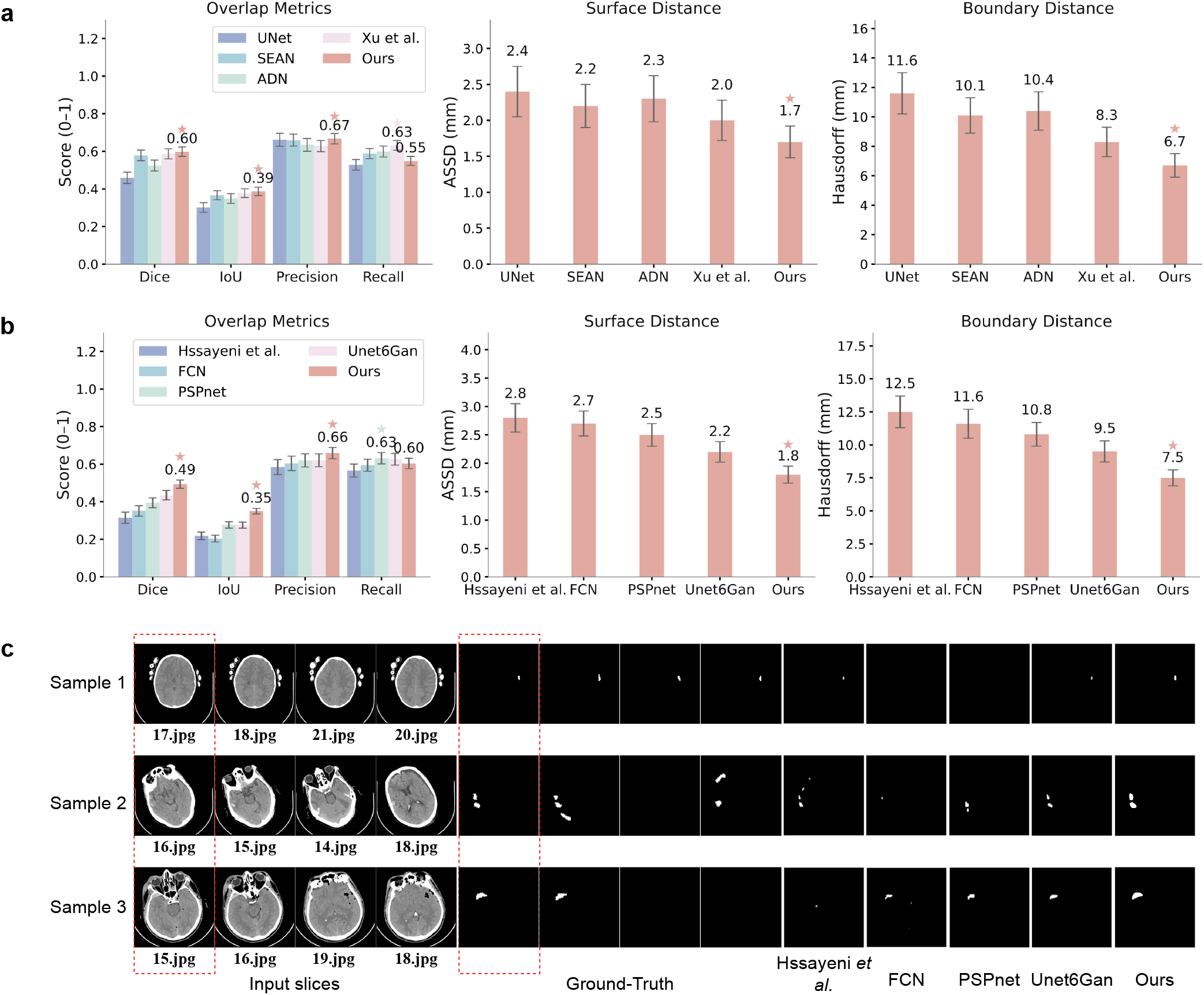
Experimental results for stroke lesion segmentation using a training-validation-testing setup, along with a comparison to state-of-the-art methods. (a) Quantitative comparison with existing state-of-the-art segmentation methods on AISD. (b) Quantitative comparison with existing state-of-the-art segmentation methods on PhysioNet-ICH. (c) Qualitative comparison with state-of-the-art models on PhysioNet-ICH. Input slices: target slices are in red box, rest slices are the *knn* of the corresponding target slice. Ground-truth: targets are in red box. Hssayeni *et al*., FCN, PSPnet, Unet6Gan: state-of-the-art methods. Ours: the proposed IISDS segmentation module.

On the PhysioNet-ICH^23^ dataset, we compare our method with Hyssayeni *et al*.^18^, FCN^28^, PSPnet^29^, and Unet6Gan^19^. Results in Figure 3b (see Supplementary Figure S3d for more details) show that StrokeGNN achieves the best Dice and IoU scores, significantly outperforming all competing methods. Additionally, qualitative comparisons in Figure 3e demonstrate that StrokeGNN produces more precise segmentations across multiple test cases. Notably, for a sample with tiny lesion, StrokeGNN effectively integrates contextual information from adjacent slices via its GNN module, leading to accurate detection. It also generates clear and well-bounded segmentations for larger lesions in other test cases.

These results confirm that IISDS achieves strong performance in both objective metrics and subjective evaluation, supporting its suitability for real-world stroke diagnosis tasks where lesion variability and volume must be robustly captured.

### Stroke lesion volume estimation from segmentation

To assess stroke severity, we adopt lesion volume as the primary quantitative proxy, using the same un-augmented CT scans employed in the segmentation experiments (Figure 2a). Patients without evidence of stroke are reliably filtered by the classification model of IISDS; therefore, severity estimation is performed only for cases diagnosed with ischemic or hemorrhagic stroke.

Lesion volumes are stratified into four groups using quartile thresholds (Q1–Q4), resulting in equal-sized cohorts that span the entire sample. Although this approach does not rely on clinical labels, the resulting strata naturally align with increasing neurological impairment and offer a preliminary approximation of NIHSS severity levels. Figure 4a-c and Figure 4d-f illustrate experimental results for lesion volume estimation. Top panels show the distribution of lesion volumes within these quartile groups for both ischemic and hemorrhagic stroke populations, revealing a clear monotonic progression with increasing lesion burden. To support qualitative interpretation, middle and bottom panels provide representative 2D slices and 3D reconstructions from each group. These examples illustrate the transition from small, well-circumscribed infarcts or hematomas in Q1 to large, multi-regional or diffuse lesions in Q4.

**Figure 4.**
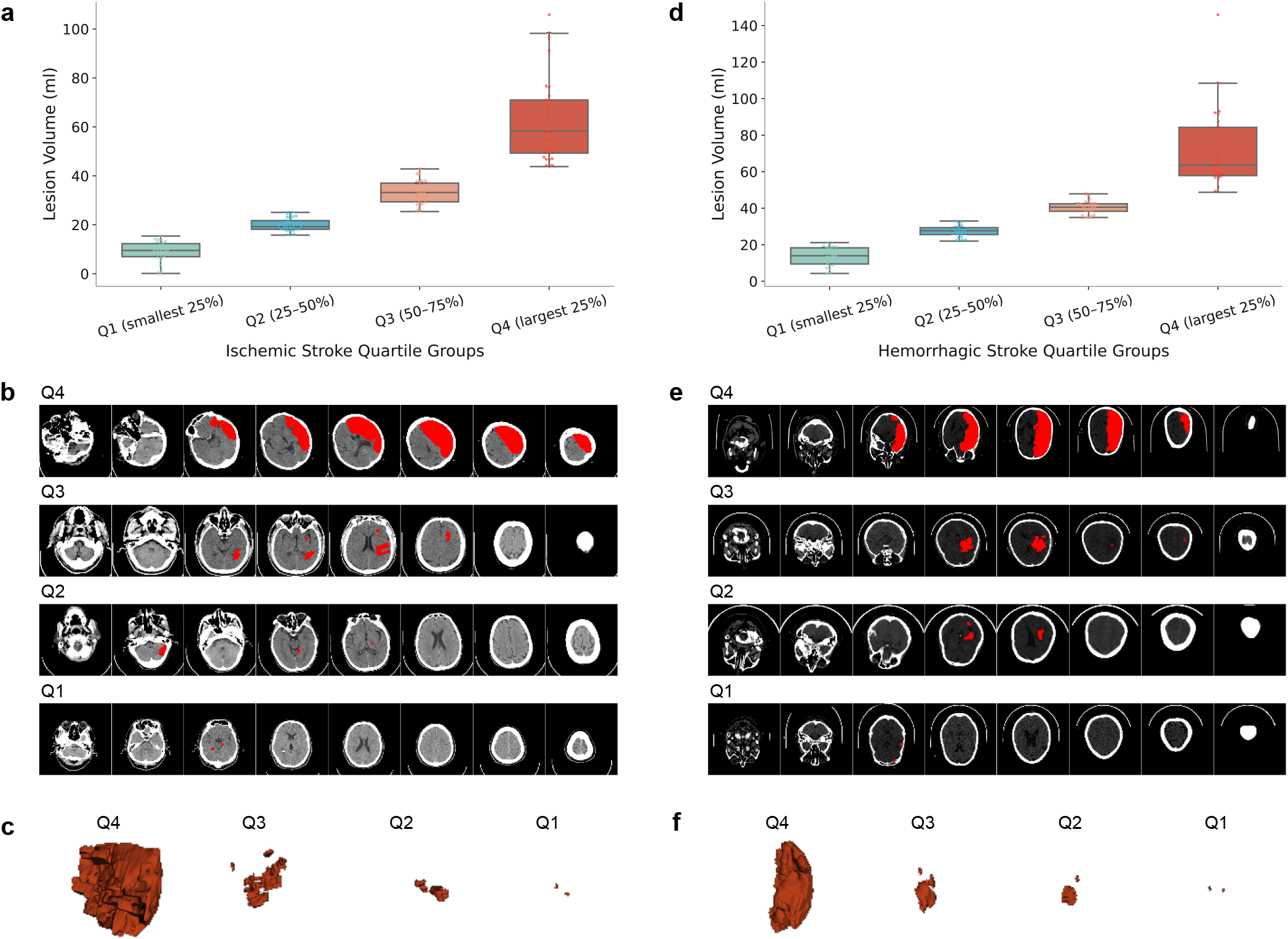
Experimental results for stroke lesion volume estimation. (a)-(c) Lesion volume groups for ischemic stroke. (a) box plot of each cluster in terms of the segmented lesion volumes, (b) sample CT scans and (c) corresponding 3D visualization of Q1-Q4 lesion volume groups (segmented lesion is highlighted in red color). (d)-(f) Lesion volume groups for hemorrhagic stroke. (d) box plot of each cluster in terms of the segmented lesion volumes, (e) sample CT scans and (f) corresponding 3D visualization of Q1-Q4 lesion volume groups (segmented lesion is highlighted in red color).

Overall, this quartile-based stratification offers a simple, interpretable, and label-free proxy for estimating NIHSS-like severity categories directly from CT imaging. While the approach is preliminary, it provides a foundation for integrating severity estimation into automated stroke assessment pipelines. Future work will incorporate additional geometric and spatial descriptors (e.g., boundary irregularity, shape asymmetry, and multi-lobar involvement), and will use clinically annotated NIHSS scores to train and validate a supervised severity-prediction model with stronger clinical fidelity.

### IISDS-enabled stroke subtype classification

As part of IISDS, stroke classification is performed as a downstream diagnostic task that leverages lesion features derived from the segmentation module. We incorporate public datasets including AISD, PhysioNet-ICH, RSNA^24^, and CQ500^25^. Notably, the RSNA^24^ dataset, the source of BHSD, includes 752,803 CT slices across 18,938 scans, annotated at the slice level by 60 members of the American Society of Neuroradiology (ASNR). CQ500^25^, the source of Seg-CQ500, contains 491 CT scans labeled at the scan level by three experienced neuroradiologists. Details can be found in Supplementary Table S6.

We first evaluate performance using five-fold cross-validation. To construct a balanced dataset, we combine AISD with 148 normal cases from RSNA to form AISD+. Data augmentation is applied to the training set to equalize class distribution across the three stroke categories. Detailed data distribution is illustrated in Figure 6a (see Supplementary Figure S4a for details). For each fold, 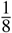 of the training set is reserved for validation. Evaluation metrics include AUC, accuracy, precision, recall, specificity, and F1-score. Training stops at approximately 33 epochs (150 minutes on average). Figure S4b illustrates the convergence behavior; optimal validation accuracy is achieved at the 26^*th*^ epoch, and training is stopped at the 36^*th*^ epoch.

Quantitative results across four datasets are summarized in Figure 6c (see Supplementary Figure S4c for details). The proposed model achieves AUC scores of 0.9600 for ischemic stroke and 0.9319 for hemorrhagic stroke. Dataset-specific AUCs include 0.9625 (AISD+), 0.8393 (PhysioNet-ICH), 0.9457 (RSNA), and 0.9481 (CQ500). Except for PhysioNet-ICH^15^, which contains only 16 annotated samples, all datasets report AUCs exceeding 0.94, underscoring the model’s generalization. False positive and false negative examples are visualized in Figure 6d, highlighting model limitations in detecting subtle lesions or class boundary ambiguities.

To enable reliable development and evaluation of stroke classification models, we establish the BGD-ISD dataset, a multi-center CT collection sourced from six major medical institutions across Bangladesh: Asgar Ali Hospital Ltd; City Hospital Ltd, Lalmatia; Ideal CT Scan Centre; International Medical College and Hospital; Popular Diagnostic Centre Ltd, Jatrabari Branch; and Popular Diagnostic Centre Ltd, Savar. The dataset contains 1,507 CT scans from 597 patients acquired between July 10^*th*^, 2023 and June 14 ^*th*^, 2024. All scans were independently annotated by two certified radiologists with clinical expertise in stroke. Detailed demographic information is provided in Figure 5 (visual) and Table S1 (numerical). Importantly, BGD-ISD represents the first large-scale, clinically annotated CT stroke dataset from Bangladesh, providing essential diversity in scanner types, clinical settings, and patient populations, filling a critical gap in global stroke imaging resources.

**Figure 5.**
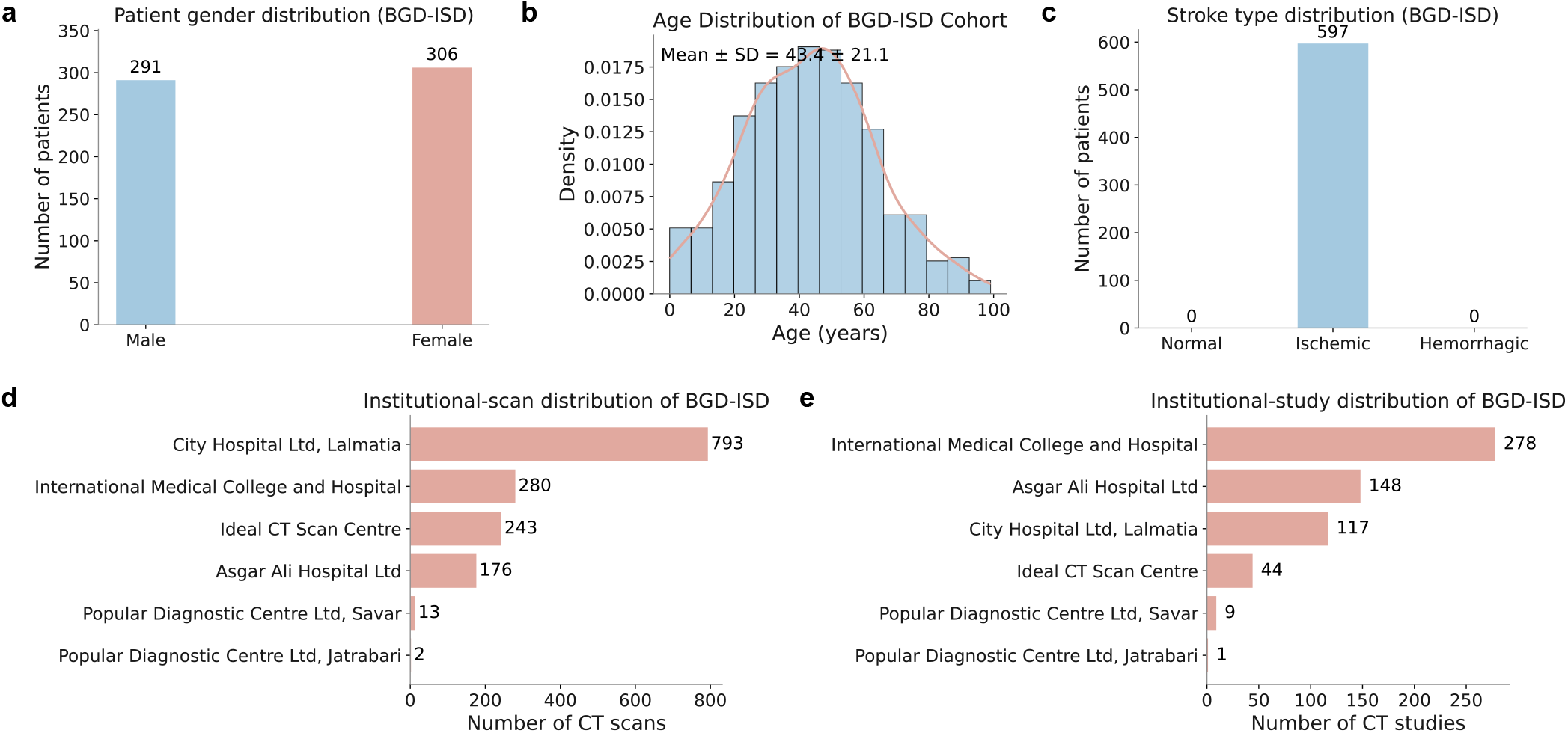
Demographic overview of the BGD-ISDcohort. (a) Gender distribution of the enrolled subjects. (b) Age distribution of patients, shown as a histogram with density estimation (mean ± s.d. indicated). (c)Distribution of stroke types across the cohort. (d) Institutional distribution by number of studies contributed from each participating medical centre. (e) Institutional distribution by number of CT scans, reflecting inter-site variability in imaging volume.

**Figure 6.**
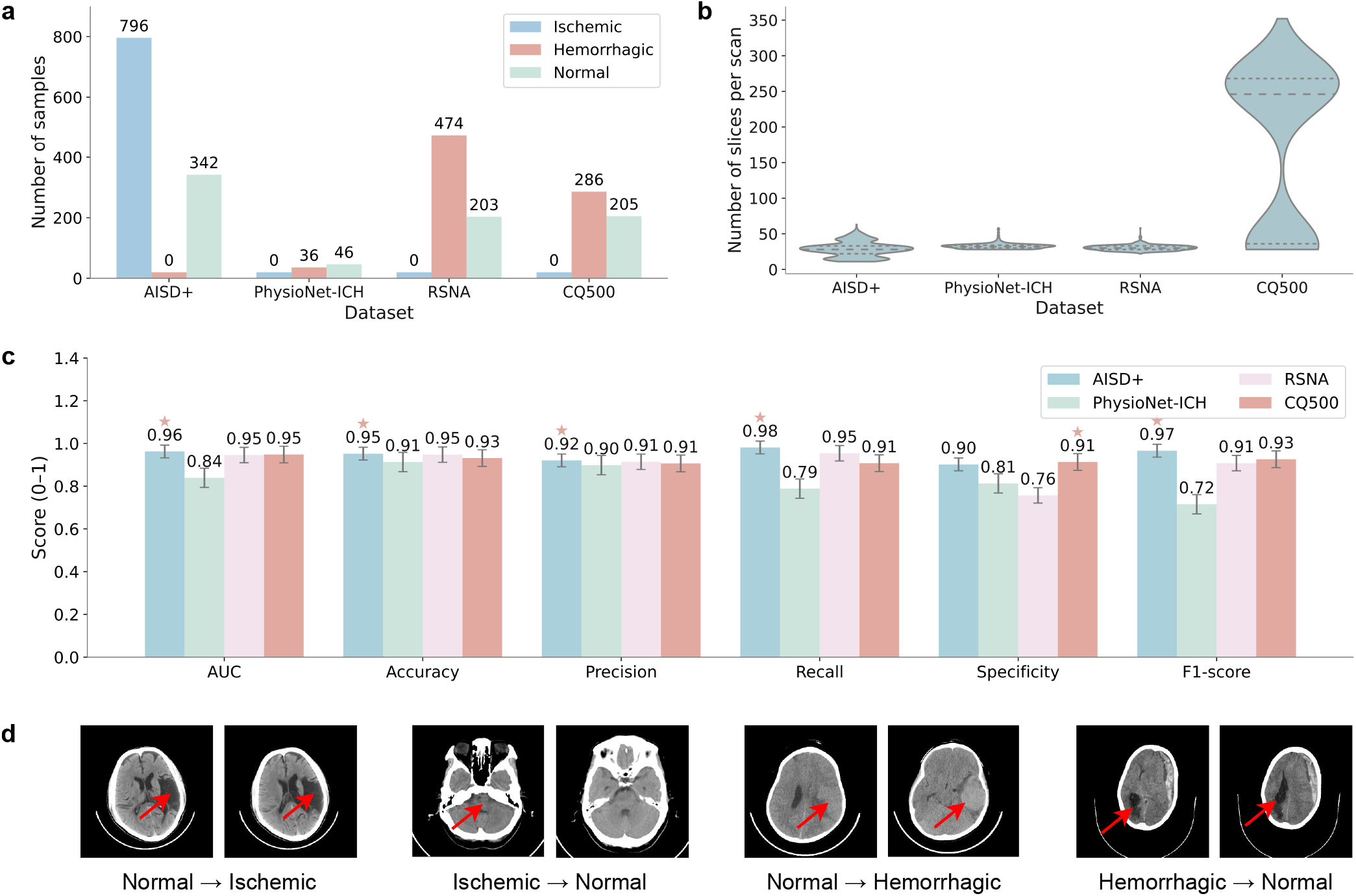
Experimental results for stroke subtype classification using a cross-validation setup. (a) Dataset distribution. (b) Slice count (z-axis) distribution for each dataset. (c) Quantitative testing results including AUC, accuracy, precision, recall, specificity, and F1-score across the original four datasets. (d) False positive and false negative samples of ischemic and hemorrhagic stroke.

To train the stroke classification framework, we use 796 stroke and 796 normal cases from RSNA together with augmented AISD data, reserving one-eighth for validation. Model convergence is achieved after 42 epochs (310 minutes), as illustrated in Figure S5b. We then benchmark the proposed method on two evaluation settings: (1) external validation on CQ500, and (2) comparative evaluation on our newly collected BGD-ISD dataset.

For external evaluation on the CQ500 dataset^11^, we compare our model against widely adopted baselines, including Chilamkurthy *et al*.^11^, ResNet^13^, SE-ResNeXt^30^, and EfficientNet-B0^14^. As shown in Table 7a (see Figure S5d for details), our approach achieves the best performance across all evaluation metrics. More importantly, unlike these prior works, our framework supports multi-class classification of ischemic and hemorrhagic stroke, demonstrating both improved accuracy and enhanced clinical utility. Consistent with these results, the ROC curves in Figure 3c further confirm the improved voxel-level discrimination capability of our model across a wide range of operating points.

On the BGD-ISD dataset, we evaluate against state-of-the-art 3D CT classification models, including Zunair *et al*.^31^, Solovyev *et al*.^32^, and Pecoraro *et al*.^33^. The results, summarized in Figure 7b (see Supplementary Figure S5c for details), show that our method consistently outperforms competing approaches in precision, specificity, and F1-score. Notably, our model improves AUC by at least 0.016 compared with other methods, further confirming the robustness of our design and the value of the newly collected BGD-ISD dataset as a new real-world benchmark. This performance advantage is further supported by ROC analysis in Figure 3d, where StrokeGNN consistently attains the highest AUC.

**Figure 7.**
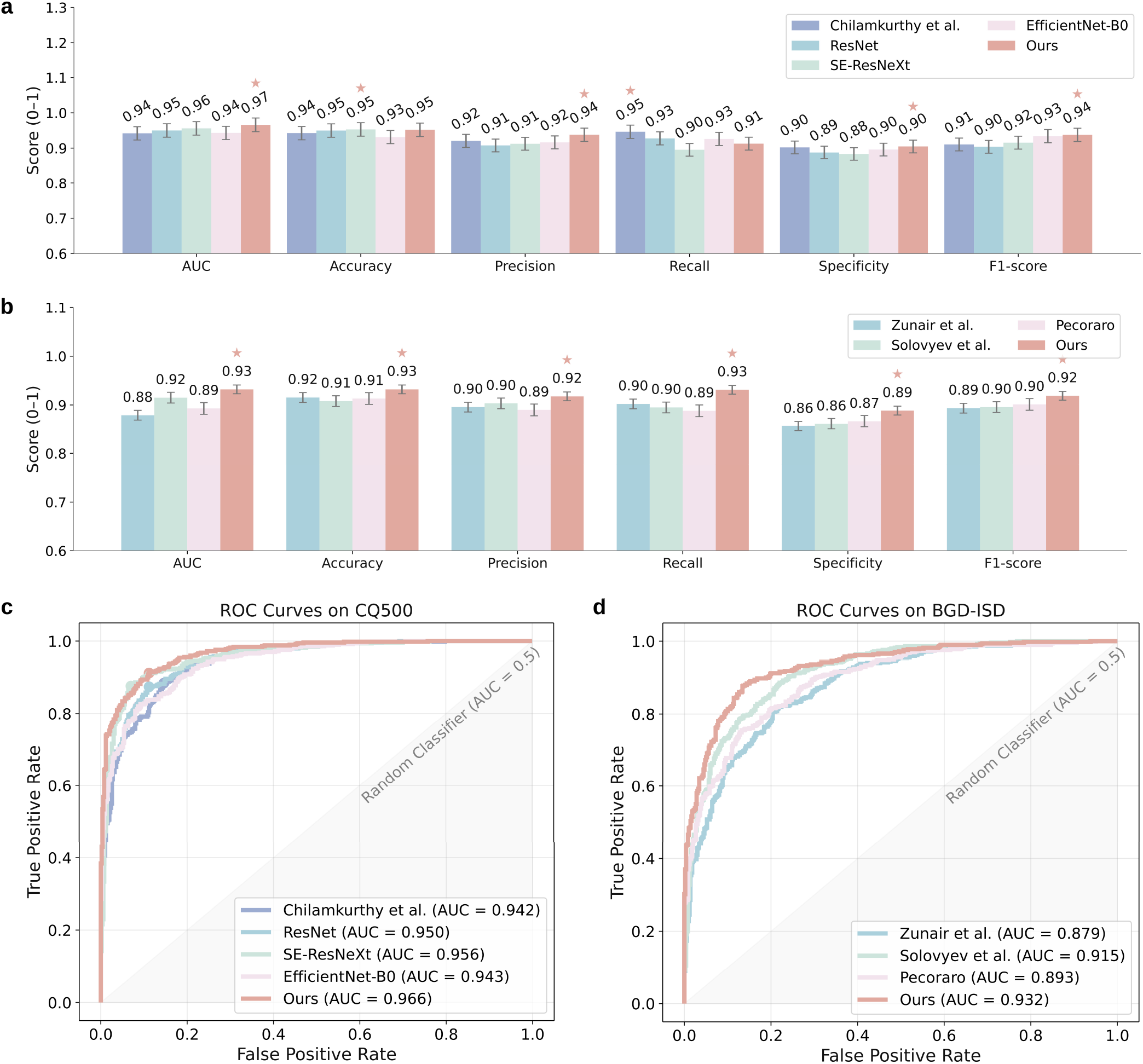
Experimental results for stroke subtype classification using a training-validation-testing setup, with a comparison to state-of-the-art methods. (a) Quantitative comparison with existing state-of-the-art methods on CQ500. (b) Quantitative comparison with existing state-of-the-art methods on the curated BGD-ISD. (c)(d) ROC curves for above CQ500 and BGD-ISD.

### Ablation study

We conducted a series of ablation experiments to evaluate the design choices that shape the performance of StrokeGNN across segmentation and classification tasks. These experiments isolate the contributions of core architectural components, graph construction strategies, and training configurations.

For the segmentation module, we first compared StrokeGNN against simple 2D and 3D U-Net backbones. As shown in Supplementary Table S3a, our design substantially improves Dice, IOU, precision, recall, and surface-distance metrics, confirming the benefit of modelling volumetric features and relationships through graph-based design. We then examined graph neighborhood size *k*, as reported in Table S3b. Performance peaks at *k* = 3, with a larger number of neighborhoods leading to oversmoothing and decreased accuracy. Both ablation studies reveal the importance of inter-slice relationships. Finally, Table S3c demonstrates that the two-head configuration (separately modelling ischemic and hemorrhagic) significantly enhances Dice scores for both stroke types compared with a single shared head. Together, these studies illustrate that both the graph topology and multi-head design contribute significantly to accurate lesion localisation segmentation.

For the classification module, Supplementary Table S4a compares 2D and 3D CNN baselines. The 3D variant achieves higher AUC, accuracy, recall, and F1-score, confirming the necessity of modelling full volumetric context for stroke subtype determination. We further evaluated the effect of transfer learning (Supplementary Table S4b), where a pre-trained model on the segmentation task provides consistent gains across all metrics. This highlights the importance of localizing lesion territories for classifying stroke subtypes.

Overall, the ablation results validate StrokeGNN’s core architectural decisions, including volumetric reasoning, optimised graph neighbourhoods, multi-head lesion modelling, and transfer-learning–based classification.

## Discussion

AI-based stroke diagnosis has demonstrated great potential in improving patient outcomes by enabling rapid localization, classification, and treatment prioritization. In this work, we present IISDS, a comprehensive stroke diagnostic framework that integrates stroke lesion segmentation, type classification, and severity assessment using only non-contrast CT scans. To the best of our knowledge, this is the first study to integrate these three key components into an end-to-end system, forming a clinical tool capable of supporting early diagnosis and decision-making. This integration aligns closely with real-world diagnostic workflows, where time-sensitive decisions must be made based on initial CT imaging in the absence of other modalities such as magnetic resonance imaging (MRI)^34^.

Lesion segmentation plays a fundamental role in stroke diagnosis, as it helps localize infarcted or hemorrhagic regions that guide further treatment. Most existing approaches rely on slice-wise 2D models or volumetric 3D convolutional neural networks that lack explicit spatial structure. The segmentation module in StrokeGNN introduces novelty by combining a standard U-Net with a 3D GNN that captures inter-slice relationships. This design allows the model to leverage both local pixel-wise features and long-range anatomical context. Our evaluation across four public datasets demonstrates consistent performance improvements over baselines, with the highest Dice score and precision observed on average. Clinically, such accurate segmentation is essential not only for estimating lesion extent but also for feeding downstream tasks like severity assessment.

Stroke volume estimation adds an additional layer of functionality by prioritizing care for patients at greater risk. Given the lack of scale labels such as NIHSS in most public datasets, we propose an unsupervised clustering strategy that estimates severity from lesion geometry information alone. The novelty here lies in the model’s ability to classify stroke patients into four clinically interpretable categories — minor, moderate, moderate to severe, and severe. Visual results of cluster distributions suggest reasonable inter-class separation. While this component remains under-explored, it highlights the potential for imaging-derived severity estimation, which could be particularly valuable in real-world settings.

Stroke type classification is critical for determining immediate intervention. For example, in acute ischemic stroke, thrombolytics therapy can restore blood flow and improve outcomes, but if a stroke is hemorrhagic, thrombolytics is contraindicated because it may exacerbate bleeding. In hemorrhagic stroke, selected patients (for example, with large intracerebral hematomas or hydrocephalus) may undergo neurosurgical evacuation as part of management^35^. While existing models generally focus on binary classification, our StrokeGNN classification module supports multi-class diagnosis among ischemic, hemorrhagic, and no-stroke categories. This is achieved by transferring learned volumetric representations from the segmentation backbone, enhancing lesion geometry information without re-training. The classification module achieved AUCs exceeding 0.94 across four independent datasets, showing generalization across imaging protocols and institutions. From a clinical perspective, this model could function as an AI-powered assistant in emergency departments, providing stroke differentiation where neuroradiologists may not be immediately available.

Our system is designed to be modular, interpretable, and compatible with existing radiology workflows. Each sub-module can operate independently or in combination, enabling integration into real-world clinical decision support platforms. For example, segmentation maps could be embedded directly onto radiological viewers; volume estimation could guide immediate assignments or transfer decisions; and classification outputs could prompt appropriate treatment. The ability to operate using only CT scans is particularly significant, given that CT is the first-line imaging modality in stroke diagnosis worldwide, especially in rural hospitals. Supplementary Figures S6-S9 show representative samples with doctor-level diagnostic quality, particularly in lesion segmentation. Supplementary Figure S10 shows a representative sample from our collected BGD-ISD, demonstrating the model’s ability to generalize to data from a different medical center..

Nonetheless, several limitations remain that should be addressed in future work. First, the lack of annotated severity labels limits the interpretability and benchmarking of the severity assessment module. Second, while the Bangladesh dataset introduces valuable diversity, broader validation will be essential to confirm the generalizability of our framework. Third, although the system shows strong quantitative and qualitative performance, its clinical utility must ultimately be validated in trials and real-world workflows. Failure cases are shown in Figure 6d and Supplementary Figures S11 and S12, highlighting that the proposed model may underperform when the images contain substantial noise or when lesions are very tiny.

Future directions include expanding annotated datasets, and further integrating with large language models (LLMs) to generate full radiology reports. We also plan to incorporate the severity assessment framework against NIHSS scores and explore regulatory pathways for deployment. Overall, IISDS offers a scalable, explainable, and clinically grounded solution for comprehensive stroke diagnosis using CT imaging. Its design, strong empirical performance, and alignment with clinical needs make it a promising step toward AI-assisted stroke care in different settings worldwide.

## Methods

To enable a comprehensive and timely stroke diagnosis from imaging data, we propose IISDS, an end-to-end system that takes patient CT scans as input and produces lesion segmentation, stroke type classification, and severity prediction. To train and evaluate IISDS, we utilize a combination of public and private datasets to ensure both diversity and feasibility. Once trained, the system is designed to deliver accurate and prompt diagnostic outputs suitable for deployment in clinical environments, including hospitals and emergency care settings. This section outlines the architecture of IISDS, including the design of the StrokeGNN module and its integration across the three core diagnostic tasks. Detailed model implementation specifications are provided in Supplementary Table S2.

### StrokeGNN layer

Images are usually treated as a grid in traditional CNN^36^, where 3D features are extracted by a 3D-Conv layer. However, due to the nature of CNNs, extracted features from border slices might be inaccurate if we want to preserve the input size and apply a padding technique. This issue is significant if border slices contain essential stroke information. Conversely, geometric deep learning techniques offer a more flexible structure for connected and complex objects. GNN was introduced by Scarselli *et al*.^37^, based on the advantages of embedding a graph structure directly within the learning process. In this paper, inspired by Vision GNN proposed by Han *et al*.^38^, we design our own StrokeGNN layer as illustrated in Figure S1a. The input feature map from U-Net has size 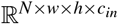, where *N* stands for the number of CT slices of a sample, *w*×*h* is the spatial dimensions, and *c*_*in*_ represents the number of input channels. The idea behind our approach is that these slices can be viewed as a set of nordered nodes in the graph as *V* = {*v*_1_, *v*_2_, …, *v*_*N*_ }. For each node *v*_*i*_ ∈*V* , its *k* (we set *k* = 3 in this paper) nearest neighbors *K* (*v*_*i*_) will be found, and edges *e* _*j,i*_ will be established from node *v* _*j*_ to node *v*_*i*_ for all *v* _*j*_ ∈ *K* (*v*_*i*_). *E* = *e*_1,1_, *e*_2,1_, …, *e*_*k,N*_ denotes directed edges in the graph, by combining with nodes *V* we obtain graph *G* = (*V, E*). To explicitly define the graph construction, for each node *v*_*i*_, its neighbourhood is defined as:

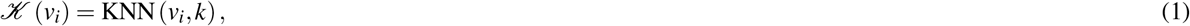

where KNN denotes the *k*-nearest neighbours in feature space and *k* = 3 in this work. The adjacency matrix *A*∈ *{*0, 1*}* ^*N*×*N*^ is defined as:

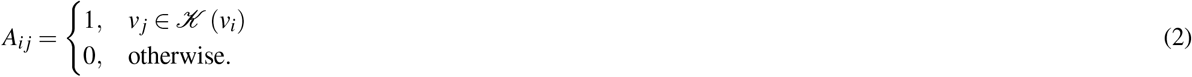

In order to extract features from each node *v*_*i*_ with size *v*_*i*_ 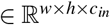and we introduce *F*(*·*) to update features of *v*_*i*_ from connected nodes:

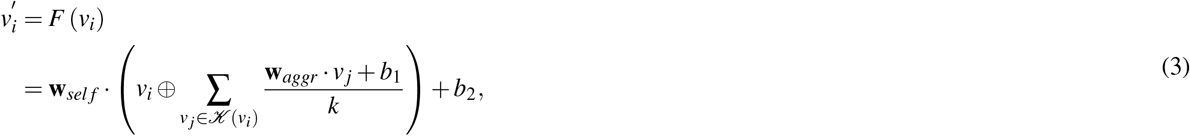

where 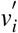 is updated features from *v*_*i*_, *⊕* stands for concatenation, **w**_*aggr*_ and **w**_*sel f*_ are learnable weights, and *b*_1_ and *b*_2_ are bias items. To be more specific, the kernel size of **w**_*aggr*_ is 3 3, together with *c*_*in*_ as both input and output channel number. The extracted feature map 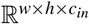is then averaged and concatenated with *v*_*i*_ to form 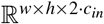features. **w**_*sel f*_ has 3 3 kernel size, 2 *c*_*in*_ input channel number, and *c*_*out*_ output channel number, yielding 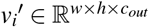 The StrokeGNN layer can also be interpreted as a message-passing operation. For each node *v*_*i*_, messages from neighbouring nodes are first aggregated as:

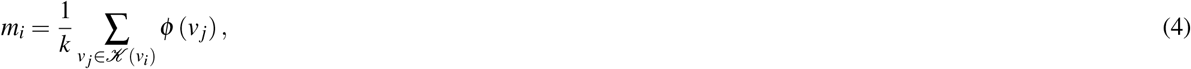

where Φ (*·*) is a learnable convolutional transformation implemented by **w**_*aggr*_. The updated node feature is then computed as:

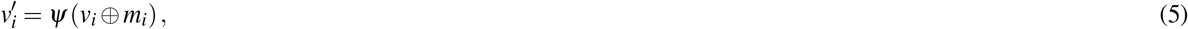

where *Ψ*(*·*) denotes a learnable transformation parameterized by **w**_*sel f*_. This formulation enables information propagation cross slices while preserving spatial resolution.

To introduce non-linearity into the StrokeGNN layer for real complex data^39^, we apply a rectified linear unit (ReLU) activation function after applying **w**_*aggr*_ and **w**_*sel f*_ as:

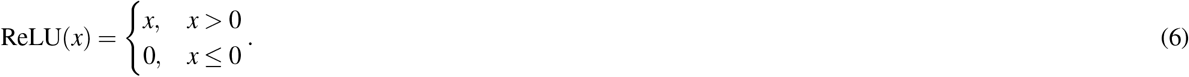

### Stroke segmentation

The stroke segmentation module adopts an enhanced U-Net architecture^27^, illustrated in blue in Figure S1a. All CT scans were converted to Hounsfield units, windowed to [100, 300] HU, and linearly normalised to [0, 1] using identical parameters across all datasets to ensure intensity consistency. They are further normalised by resampling to the same in-plane resolution. The input to the model is a pre-processed CT volume of size R*D*×*W*×*H*×1. To reduce GPU memory usage and stabilize training, each scan is partitioned into fixed-size patches of dimension R^*d*×*w*×*h*×1^, where *d < D* and *w < W*. These volumetric patches are processed independently during training and reassembled during inference (by majority voting for overlapping areas). Each patch is first passed through a 2D U-Net encoder–decoder structure with skip connections, where every block contains two 3 × 3 convolutions followed by ReLU activation. This 2D backbone extracts strong in-plane semantic features across all *w* × *h* slices, producing a feature tensor of size R^*d*×*w*×*h*×64^. To further capture 3D contextual information across the through-plane dimension, we incorporate two StrokeGNN layers that perform graph-based message passing along the *d*-dimension. This enhances the volumetric representation and yields a feature map of size R^*d*×*w*×*h*×2^ for binary segmentation.

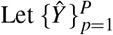 denote the predicted probability maps for all extracted patches, where overlapping regions may receive multiple predictions. During inference, the final voxel-wise prediction *Ŷ* is reconstructed by majority voting:

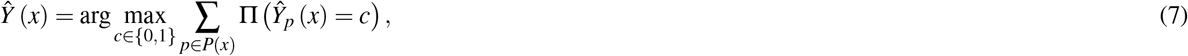

where *P*(*x*) denotes the set of patches covering voxel *x*, and Π(*·*) is the indicator function.

We adopt a dual-head prediction design, where two parallel segmentation heads specialize in ischemic and hemorrhagic lesions. Each head consists of a 1×1 convolutional layer followed by softmax activation, producing probability maps of size R^*d*×*w*×*h*×2^. This design enables the model to learn unique patterns for different lesion types. At training and testing time, annotated head is used to ensure the best model performance. At inference time, both heads are executed in parallel using the same input representation, and the final diagnosis is selected by choosing the head whose task corresponds to the predicted stroke type (ischemic or hemorrhagic). This strategy avoids additional computational overhead and ensures that the most relevant specialised head is used for the final decision.

The training loss sums the Dice loss^40^ and cross-entropy loss for each head. For ground truth *y* and predicted probabilities *p*, the loss is:

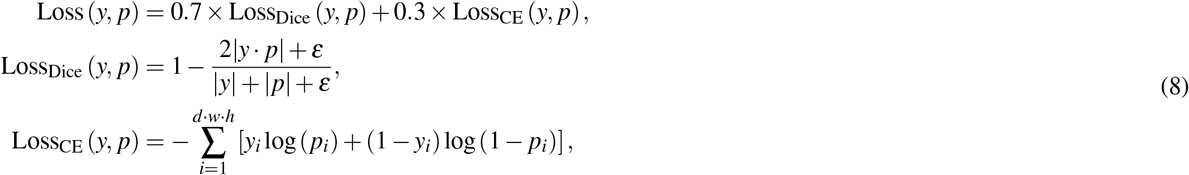

where *ε* = 1 ×10^−6^ prevents division by zero. As ArgMax is non-differentiable, training is performed on probability maps; discrete masks are used only during inference. Patient-level Dice loss is used during training, while slice-level metrics are reported at test time for comparability with prior work.

To mitigate overfitting^41^ and balance ischemic and hemorrhagic samples, we apply random augmentations including cropping within [95%, 100%] of the field of view, rotation within [−15^*°*^, 15^*°*^], and horizontal flipping with probability 0.5. Gradient accumulation is employed to achieve an effective batch size of 2.

In summary, the segmentation module combines (i) 2D U-Net for strong intra-slice representation, (ii) StrokeGNN layers for volumetric reasoning across patches, and (iii) dual ischemic/hemorrhagic heads for their corresponding segmentation task. The model operates on patches of size R^*d*×*w*×*h*×1^ and outputs lesion probability maps of the same size. Training uses the RMSprop optimizer with a learning rate of 1× 10^−5^, with early stopping applied if validation loss does not improve by more than 0.002 over five consecutive epochs.

### Stroke volume estimation

Stroke lesion volume serves as an essential quantitative biomarker for assessing stroke severity and guiding subsequent analysis tasks. Following lesion segmentation, we compute volume directly from the predicted 3D mask by summing all lesion voxels and multiplying by the physical voxel spacing. This yields an anatomically accurate estimate in millilitres (mL) without requiring additional geometric assumptions.

Formally, for a predicted binary mask *M* ∈ *{*0, 1*}*^*D*×*W*×*H*^ and voxel spacing (*s*_*d*_, *s*_*w*_, *s*_*h*_), the lesion volume is given by

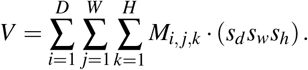

In practice, voxel spacing is obtained from the DICOM metadata, ensuring that the estimated volume remains invariant to scanner resolution and resampling operations. This discrete summation provides an exact volumetric measurement under the voxel-grid representation.

This volume estimate has substantial potential to be used for downstream severity categorisation and for analysing correlations between lesion burden and model decision patterns. Because the computation is performed directly on the same un-augmented CT scans used during segmentation, the resulting volumes remain fully consistent with both the diagnostic and multi-task learning components of our workflow.

### Stroke classification

The stroke classification task utilizes a transfer learning technique, which has been proven to reduce training times and achieve better performance than training from scratch^42^. In this paper, as shown in Figure S1a, we will take the bottleneck features from the previous stroke segmentation task to classify scans into three classes: “no stroke”, “ischemic stroke”, or “hemorrhagic stroke”. Firstly, we take 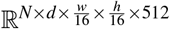 intermediate features. Secondly, we apply global averaging pooling and obtain R^*N*×512^ feature, which is formally defined as:

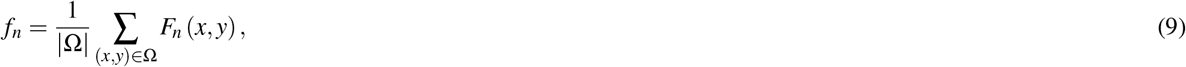

where 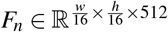 denotes the feature map of slice *n*, Ω indexes spatial location, and *f*_*n*_ ∈ R^512^ is the pooled slice-level feature. Thirdly, we apply attention pooling and obtain R^1×512^ features as:

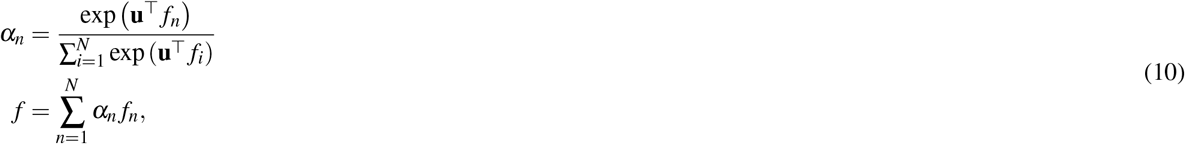

where **u** is a learnable attention vector, *α*_*n*_ denotes the attention weight for slice *n*, and *f* ∈ R^512^ is the final scan-level feature. Eventually, we apply 3 fully-connected layers (i.e., dense layers) with drop_out = 0.1 to prevent overfitting and yield R^3^ features for 3-class classification. Eventually, we apply softmax for probability transformation and cross-entropy loss as:

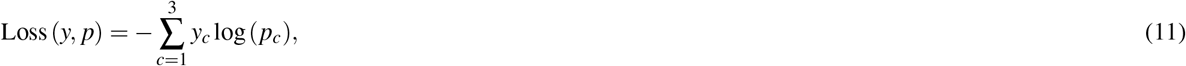

where *y* is the ground-truth class label and *p* is the predicted probabilities.

The stroke classification model leverages intermediate feature representations extracted from the segmentation network via transfer learning. Training is performed using cross-entropy loss, with the SGD optimizer (learning rate = 1 × 10^−3^, momentum = 0.9), and L2 regularization (weight decay = 0.001) to mitigate overfitting. The model is trained with a batch size of 2 and early stopping criteria is triggered if validation loss fails to improve for 10 consecutive epochs.

## Data Availability

All data produced will be available online at https://github.com/Zhicheng-Lu/stroke_ct upon the acceptance of the paper.

## Implementation details

Model training was conducted on a workstation equipped with an Intel Core i7-13700 CPU, an NVIDIA RTX 4090 GPU, and 32 GB of RAM, running Ubuntu 20.04. The proposed models were implemented in PyTorch, with GPU acceleration enabled via CUDA 11.8.

## Data availability

Link of BGD-ISD from Bangladesh we collect can be found in: https://github.com/Zhicheng-Lu/stroke_ct.

All other data in this paper are from publicly available datasets and can be downloaded from the following links (see Supplementary Table S5 and S6 for detailed overview):

- AISD: https://github.com/GriffinLiang/AISD.
- PhysioNet-ICH:https://physionet.org/content/ct-ich/1.3.1/.
- BHSD: https://huggingface.co/datasets/Wendy-Fly/BHSD.
- Seg-CQ500: https://zenodo.org/records/8063221.
- RSNA: https://www.kaggle.com/c/rsna-intracranial-hemorrhage-detection/data.
- CQ500: http://15.206.3.216/dataset.

## Code availability

All source codes are publicly available at https://github.com/Zhicheng-Lu/stroke_ct.

## Acknowledgments

This work was supported by Australian Commonwealth Funding.

## Author contributions statement

Z.Lu, S.Uddin, S.Uribe, S.White, R.T.Martins, S.Chau, A.S.M.Mosaddek, M.S.Islam, N.Nahar, A.K.Z.Azad, K.M.N.Hossain, H.S.Choudhury, K.M.R.Hasan, N.Mosaddek, S.Rahman, M.M.Hossain, and K.M.M.H.Sizar, C.Angione, P.Liò, M.T.Islam, and M.A.Moni made contributions to the concept and design of the article. Z.Lu conceived the study, designed the methodology, developed the models, conducted the experiments, and analysed the results. A.S.M.Mosaddek, M.S.Islam, N.Nahar, K.M.N.Hossain, H.S.Choudhury, K.M.R.Hasan, N.Mosaddek, S.Rahman, M.M.Hossain, and K.M.M.H.Sizar contributed to data curation and clinical interpretation. S.Uribe, S.White, R.T.Martins, and S.Chau provided domain expertise and guidance on imaging protocols. M.A.Moni supervised the project. All authors reviewed and approved the manuscript.

