## Supplementary for "Clinically Generalisable End-to-End Graph Learning for CT Image-Based Multitask Stroke Diagnosis"

#### This PDF file includes:

Figs. S1 to S12
Tables S1 to S6
SI References

### Supplementary: Demographic Information of Curated BGD-ISD

**Table S1. Demography overview of the collected BGD-ISD. (a) Patient-level characteristics, including age distribution, gender ratio, and stroke type breakdown. (b) Institutional distribution of the dataset across contributing medical centres.**

| Characteristic | Value |
|---|---|
| Total subjects | 597 |
| Age (mean $\pm$ SD) | 43.4 $\pm$ 21.1 years |
| Age range | 0 - 99 |
| Gender (M / F) | 291 (48.7%) / 306 (51.3%) |
| Stroke type | 597 ischemic (100%) |

**(a)**

| Institution | Studies | Scans |
|---|---|---|
| Asgar Ali Hospital Ltd | 148 | 176 |
| City Hospital Ltd, Lalmatia | 117 | 793 |
| Ideal CT Scan Centre | 44 | 243 |
| International Medical College and Hospital | 278 | 280 |
| Popular Diagnostic Centre Ltd, Jatrabari Branch Dhaka | 1 | 2 |
| Popular Diagnostic Centre Ltd, Savar | 9 | 13 |
| **Total** | 597 | 1,507 |

**(b)**

### Supplementary: Model Details

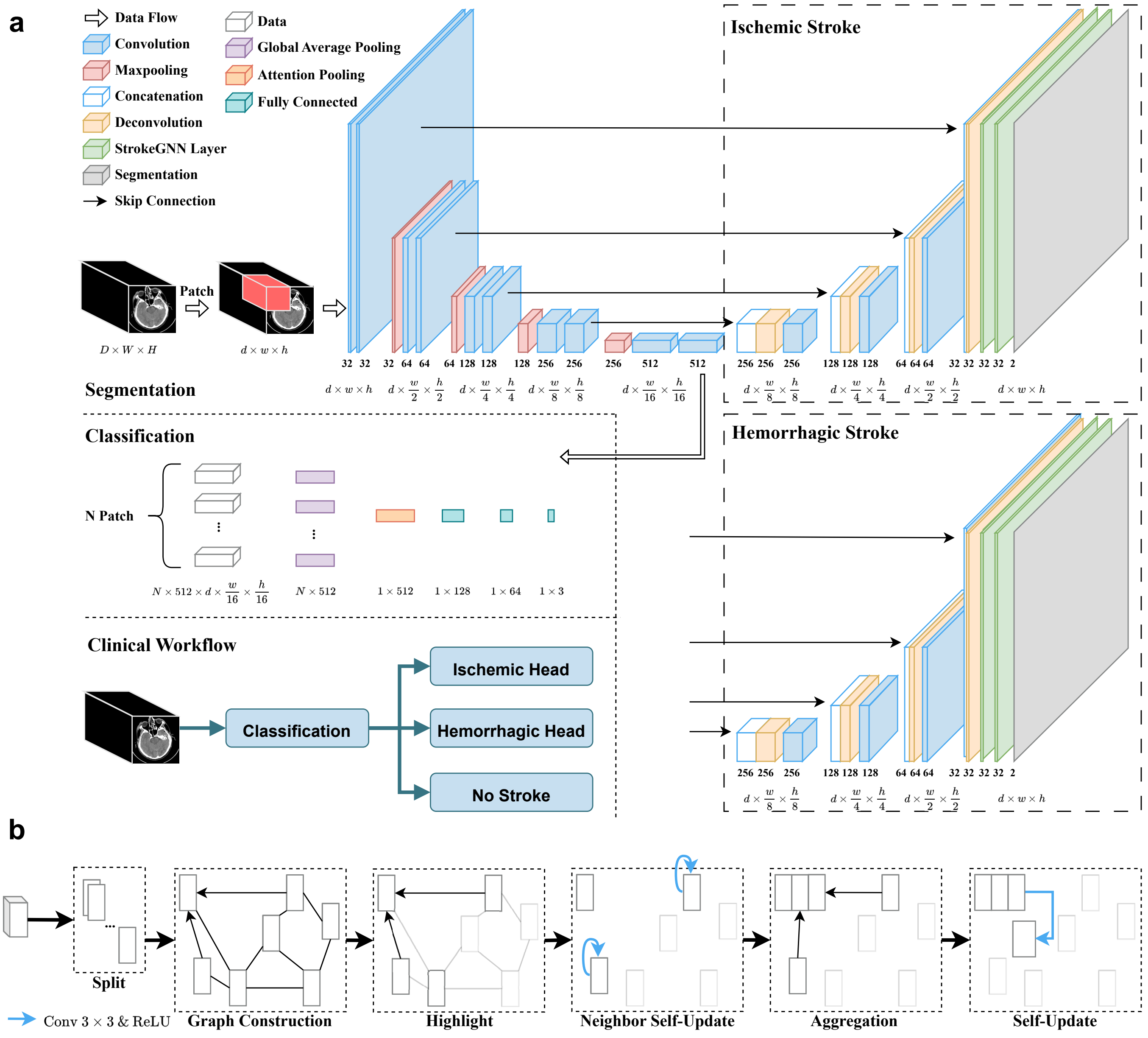


(a)

**Fig. S1.** Proposed IISDS, StrokeGNN and StrokeGNN layer. (a) Overview of the proposed IISDS. Patched 3D CT scans are fed into two-head U-Net structure to segment of ischemic and hemorrhagic strokes separately. Bottleneck features are fed into classification model to categorize stroke types. In real clinical workflow, input CT scan is classified first then fed into corresponding segmentation head. (b) Overview of StrokeGNN layer. CT scans are split into slices where each slice is regarded as a node in the graph. Edges are constructed by $K$ nearest neighbors of each node. A node will aggregate features from its neighbors and update itself for next model layer.

**Table S2. Proposed model details. (a) Stroke segmentation module details. All convolutional layers use $1\times1$ stride and "same" padding, and apply ReLU activation function afterwards except final "Conv" layer. (b) Stroke Classification module details. Input is from Table S2a Up1_2 layer. All convolutional and fully connected layers use $1\times1$ stride and "same" padding, and apply ReLU activation function afterwards except final "FC3" layer.**

| | | |
|---|---|---|
| Down1 | Down1_1 | Input: $\mathbb{R}^{1\times d\times w\times h}$. Kernel: $3\times3$. Out_channel : 32. |
| | Down1_2 | Kernel: $3\times3$. Out_channel: 32. |
| | Maxpool1 | Stride: $2\times2$. |
| Down2 | Down2_1 | Input: Maxpool1 $\in\mathbb{R}^{32\times d\times\frac{w}{2}\times\frac{h}{2}}$. Kernel: $3\times3$. Out_channel: 64. |
| | Down2_2 | Kernel: $3\times3$. Out_channel: 64. |
| | Maxpool2 | Stride: $2\times2$. |
| Down3 | Down3_1 | Input: Maxpool2 $\in\mathbb{R}^{64\times d\times\frac{w}{4}\times\frac{h}{4}}$. Kernel: $3\times3$. Out_channel: 128. |
| | Down3_2 | Kernel: $3\times3$. Out_channel: 128. |
| | Maxpool3 | Stride: $2\times2$. |
| Down4 | Down4_1 | Input: Maxpool3 $\in\mathbb{R}^{128\times d\times\frac{w}{8}\times\frac{h}{8}}$. Kernel: $3\times3$. Out_channel: 256. |
| | Down4_2 | Kernel: $3\times3$. Out_channel: 256. |
| | Maxpool4 | Stride: $2\times2$. |
| Up1 | Up1_1 | Input: Maxpool4 $\in\mathbb{R}^{256\times d\times\frac{w}{16}\times\frac{h}{16}}$. Kernel: $3\times3$. Out_channel: 512. |
| | Up1_2 | Kernel: $3\times3$. Out_channel: 512. |
| | UpConv1 | Kernel: $3\times3$. Stride: $2\times2$. Out_channel:256. |
| Up2 | Concat1 | Input: Down4_2 $\in\mathbb{R}^{256\times d\times\frac{w}{8}\times\frac{h}{8}}$, UpConv1 $\in\mathbb{R}^{256\times d\times\frac{w}{8}\times\frac{h}{8}}$. Axis: 0. |
| | Up2_1 | Kernel: $3\times3$. Out_channel: 256. |
| | Up2_2 | Kernel: $3\times3$. Out_channel: 256. |
| | UpConv2 | Kernel: $3\times3$. Stride: $2\times2$. Out_channel: 128. |
| | Concat2 | Input: Down3_2 $\in\mathbb{R}^{128\times d\times\frac{w}{4}\times\frac{h}{4}}$, UpConv2 $\in\mathbb{R}^{128\times d\times\frac{w}{4}\times\frac{h}{4}}$. Axis: 0. |

| | | |
|---|---|---|
| Up3 | Up3_1 | Kernel: $3\times3$. Out_channel: 128. |
| | Up3_2 | Kernel: $3\times3$. Out_channel: 128. |
| | UpConv3 | Kernel: $3\times3$. Stride: $2\times2$. Out_channel: 64. |
| Up4 | Concat3 | Input: Down2_2 $\in\mathbb{R}^{64\times d\times\frac{w}{2}\times\frac{h}{2}}$, UpConv3 $\in\mathbb{R}^{64\times d\times\frac{w}{2}\times\frac{h}{2}}$. Axis: 0. |
| | Up4_1 | Kernel: $3\times3$. Out_channel: 64. |
| | Up4_2 | Kernel: $3\times3$. Out_channel: 64. |
| | UpConv4 | Kernel: $3\times3$. Stride: $2\times2$. Out_channel: 32. |
| | Concat4 | Input: Down1_2 $\in\mathbb{R}^{32\times d\times w\times h}$, UpConv4 $\in\mathbb{R}^{32\times d\times w\times h}$. Axis: 0. |
| GNN1_ KNN | Build1 | Input: Concat4 $\in\mathbb{R}^{64\times d\times w\times h}$. Output: $\mathbb{R}^{3\times64\times d\times w\times h}$. |
| | Aggr1 | Kernal: $3\times3$. Out_channel: 64. |
| | Mean1 | Axis: 0. |
| GNN1 | Concat5 | Input: Concat4 $\in\mathbb{R}^{64\times d\times w\times h}$, Mean1 $\in\mathbb{R}^{64\times d\times w\times h}$. Axis: 0. |
| | Update1 | Kernel: $3\times3$. Out_channel: 64. |
| GNN2_ KNN | Build2 | Input: Update1 $\in\mathbb{R}^{64\times d\times w\times h}$. Output: $\mathbb{R}^{3\times64\times d\times w\times h}$. |
| | Aggr2 | Kernal: $3\times3$. Out_channel: 64. |
| | Mean2 | Axis: 0. |
| GNN2 | Concat6 | Input: Update1 $\in\mathbb{R}^{64\times d\times w\times h}$, Mean2 $\in\mathbb{R}^{64\times d\times w\times h}$. Axis: 0. |
| | Update2 | Kernel: $3\times3$. Out_channel: 64. |
| | Conv | Kernel: $1\times1$. Out_channel: 2. |
| | Softmax | Axis: 0. |

**(a)**

| | | |
|---|---|---|
| Average | Patches | Input: Up1_2 $\in\mathbb{R}^{512\times d\times\frac{w}{16}\times\frac{h}{16}}$. Output: $\mathbb{R}^{N\times512\times d\times\frac{w}{16}\times\frac{h}{16}}$. |
| | G_Average | Out_features: $N\times512$. |
| Attention | A_Pooling | Input: G_Average $\in\mathbb{R}^{N\times512}$. Output: $\mathbb{R}^{1\times512}$ |
| FC1 | FC1 | Input: A_Pooling $\in\mathbb{R}^{1\times512}$. Out_channel: 128. |
| | Dropout2 | p: 0.1. |
| FC2 | FC2 | Input: Dropout1 $\in\mathbb{R}^{1\times128}$. Out_channel: 164. |
| | Dropout2 | p: 0.1. |
| FC3 | FC3 | Input: Dropout2 $\in\mathbb{R}^{1\times64}$. Out_channel: 3. |
| | Softmax | Axis: 0. |

**(b)**

### Supplementary: Segmentation

| | Original | Augmented | Total |
|---|---|---|---|
| AISD (1) | 398 | - | 398 |
| PhysioNet-ICH (2) | 36 | 72 | 108 |
| BHSD (3) | 192 | - | 192 |
| Seg-CQ500 (4) | 51 | 51 | 102 |
| **Total** | 677 | 123 | 800 |

**(a)**

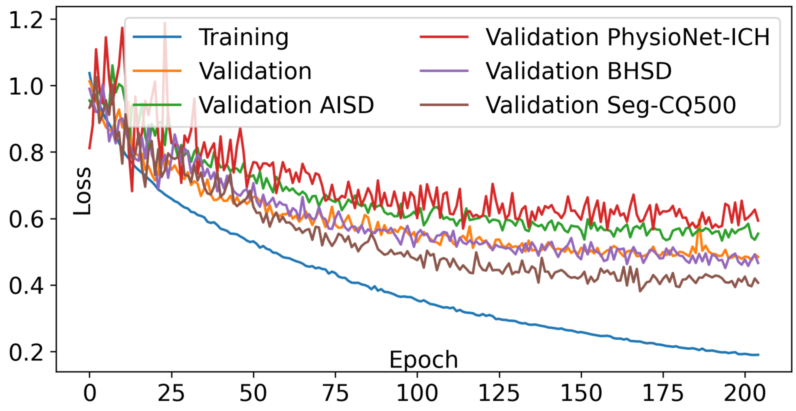


**(b)**

| | Dice ↑ | IoU ↑ | Precision ↑ | Recall ↑ | ASSD ↓ | Hausdorff ↓ |
|---|---|---|---|---|---|---|
| AISD | $0.5793 \pm 0.043$ | $0.3848 \pm 0.032$ | $0.6616 \pm 0.051$ | $0.5282 \pm 0.047$ | $1.8 \pm 0.4$ | $8.0 \pm 1.2$ |
| PhysioNet-ICH | $0.5060 \pm 0.046$ | $0.3562 \pm 0.029$ | $\mathbf{0.7750 \pm 0.058}$ | $0.4783 \pm 0.052$ | $2.2 \pm 0.5$ | $11.3 \pm 1.6$ |
| BHSD | $0.6406 \pm 0.041$ | $0.4712 \pm 0.035$ | $0.7321 \pm 0.049$ | $0.5977 \pm 0.044$ | $\mathbf{1.4 \pm 0.3}$ | $\mathbf{6.5 \pm 1.1}$ |
| Seg-CQ500 | $\mathbf{0.6910 \pm 0.039}$ | $\mathbf{0.4783 \pm 0.033}$ | $0.7027 \pm 0.042$ | $\mathbf{0.6796 \pm 0.038}$ | $1.7 \pm 0.4$ | $7.2 \pm 1.3$ |
| **Overall** | $0.6434 \pm 0.030$ | $0.4507 \pm 0.025$ | $0.7053 \pm 0.039$ | $0.5983 \pm 0.031$ | $1.8 \pm 0.2$ | $7.8 \pm 0.8$ |

**(c)**

**Fig. S2.** Experimental results for stroke lesion segmentation in 5-fold cross validation. (a) Dataset distributions before and after augmentation. (b) Training and validation losses by epochs. (c) Quantitative testing results including Dice, IoU, precision, recall, ASSD, and Hausdorff distance across the original four datasets.

| | Training (original) | Training (total) | Inference |
|---|---|---|---|
| AISD | 345 | 345 | 52 |
| PhysioNet-ICH | - | - | 36 |
| BHSD | 192 | 192 | - |
| Seg-CQ500 | 51 | 102 | - |
| **Total** | 588 | 639 | 88 |

**(a)**

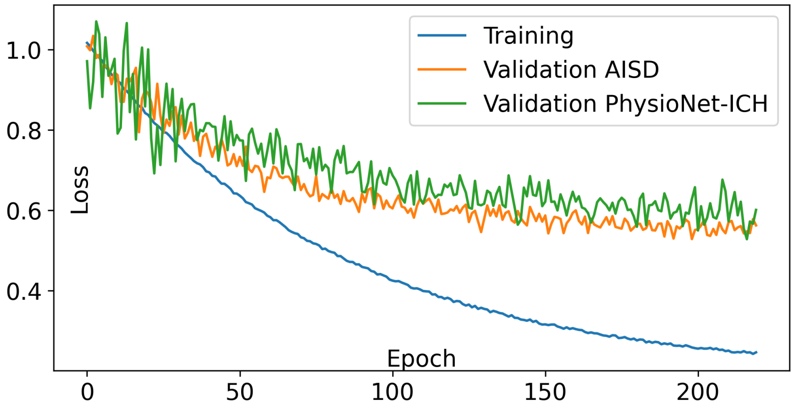


**(b)**

| | Dice ↑ | IoU ↑ | Precision ↑ | Recall ↑ | ASSD ↓ | HD ↓ |
|---|---|---|---|---|---|---|
| UNet (5) | $0.4588 \pm 0.045$ | $0.3016 \pm 0.035$ | $0.6616 \pm 0.050$ | $0.5282 \pm 0.047$ | $2.4 \pm 0.5$ | $11.6 \pm 1.9$ |
| SEAN (1) | $0.5784 \pm 0.042$ | $0.3662 \pm 0.033$ | $0.6597 \pm 0.048$ | $0.5880 \pm 0.045$ | $2.2 \pm 0.4$ | $10.1 \pm 1.6$ |
| ADN (6) | $0.5242 \pm 0.040$ | $0.3489 \pm 0.031$ | $0.6345 \pm 0.045$ | $0.5989 \pm 0.044$ | $2.3 \pm 0.4$ | $10.4 \pm 1.5$ |
| Xu et al. (7) | $0.5866 \pm 0.038$ | $0.3776 \pm 0.030$ | $0.6278 \pm 0.044$ | $\mathbf{0.6319 \pm 0.041}$ | $2.0 \pm 0.3$ | $8.3 \pm 1.4$ |
| **Ours** | $\mathbf{0.5979 \pm 0.032}$ | $\mathbf{0.3872 \pm 0.028}$ | $\mathbf{0.6673 \pm 0.040}$ | $0.5489 \pm 0.039$ | $\mathbf{1.7 \pm 0.3}$ | $\mathbf{6.7 \pm 1.2}$ |

**(c)**

| | Dice ↑ | IoU ↑ | Precision ↑ | Recall ↑ | ASSD ↓ | HD ↓ |
|---|---|---|---|---|---|---|
| Hssayeni et al. (8) | $0.3150 \pm 0.032$ | $0.2180 \pm 0.028$ | $0.5844 \pm 0.041$ | $0.5660 \pm 0.038$ | $2.8 \pm 0.4$ | $12.5 \pm 1.3$ |
| FCN (9) | $0.3512 \pm 0.034$ | $0.2038 \pm 0.025$ | $0.6043 \pm 0.045$ | $0.5945 \pm 0.041$ | $2.7 \pm 0.3$ | $11.6 \pm 1.1$ |
| PSPNet (10) | $0.3942 \pm 0.030$ | $0.2766 \pm 0.031$ | $0.6199 \pm 0.044$ | $\mathbf{0.6317 \pm 0.039}$ | $2.5 \pm 0.3$ | $10.8 \pm 1.0$ |
| UNet6-GAN (11) | $0.4348 \pm 0.028$ | $0.2754 \pm 0.027$ | $0.6206 \pm 0.037$ | $0.6262 \pm 0.041$ | $2.2 \pm 0.3$ | $9.5 \pm 0.9$ |
| **Ours** | $\mathbf{0.4937 \pm 0.029}$ | $\mathbf{0.3501 \pm 0.034}$ | $\mathbf{0.6586 \pm 0.038}$ | $0.6036 \pm 0.037$ | $\mathbf{1.8 \pm 0.2}$ | $\mathbf{7.5 \pm 0.7}$ |

**(d)**

**Fig. S3.** Experimental results for stroke lesion segmentation for comparison with state-of-the-art methods. (a) Dataset distributions before and after augmentation. (b) Training and validation losses by epochs. (c) Quantitative testing results including Dice, IoU, precision, recall, ASSD, and Hausdorff distance on AISD. (d) Quantitative testing results including Dice, IoU, precision, recall, ASSD, and Hausdorff distance on PhysioNet-ICH.

### Supplementary: Classification

| | AIS | ICH | Normal | Total |
|---|---|---|---|---|
| AISD+ (1) | 796 | - | 342 | 1138 |
| PhysioNet-ICH | - | 36 | 46 | 82 |
| RSNA (12) | - | 474 | 203 | 677 |
| CQ500 (13) | - | 286 | 205 | 491 |
| **Total** | 796 | 796 | 796 | 2388 |

**(a)**

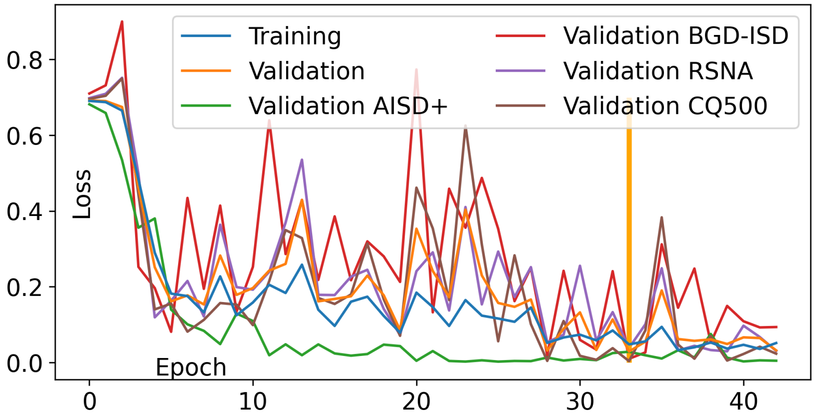


**(b)**

| | AUC ↑ | Accuracy ↑ | Precision ↑ | Recall ↑ | Specificity ↑ | F1-score ↑ |
|---|---|---|---|---|---|---|
| AISD+ | $\mathbf{0.9625 \pm 0.010}$ | $\mathbf{0.9522 \pm 0.010}$ | $\mathbf{0.9207 \pm 0.009}$ | $\mathbf{0.9812 \pm 0.012}$ | $\mathbf{0.9020 \pm 0.009}$ | $\mathbf{0.9660 \pm 0.011}$ |
| PhysioNet-ICH | $0.8393 \pm 0.017$ | $0.9130 \pm 0.013$ | $0.8986 \pm 0.014$ | $0.7886 \pm 0.018$ | $0.8129 \pm 0.020$ | $0.7154 \pm 0.019$ |
| RSNA | $0.9457 \pm 0.011$ | $0.9477 \pm 0.010$ | $0.9144 \pm 0.013$ | $0.9542 \pm 0.010$ | $0.7571 \pm 0.017$ | $0.9079 \pm 0.011$ |
| CQ500 | $0.9481 \pm 0.010$ | $0.9312 \pm 0.011$ | $0.9066 \pm 0.012$ | $0.9077 \pm 0.012$ | $0.9132 \pm 0.011$ | $0.9258 \pm 0.010$ |
| **Overall** | $0.9522 \pm 0.013$ | $0.9468 \pm 0.011$ | $0.9176 \pm 0.15$ | $0.9489 \pm 0.014$ | $0.8866 \pm 0.013$ | $0.9348 \pm 0.010$ |

**(c)**

**Fig. S4.** Experimental results for stroke subtype classification in 5-fold cross validation. (a) Dataset distributions for different categories. (b) Training and validation losses by epochs. (c) Quantitative testing results including AUC, accuracy, precision, recall, specificity, and F1-score across the original four datasets.

| | Training | | | Testing | | |
|---|---|---|---|---|---|---|
| | AIS | ICH | Normal | AIS | ICH | Normal |
| AISD | 796 | - | - | - | - | - |
| RSNA | - | 796 | 796 | - | - | - |
| CQ500 | - | - | - | - | 286 | 205 |
| BGD-ISD | - | - | - | 1507 | - | - |
| **Total** | 796 | 796 | 796 | 1507 | 286 | 205 |

**(a)**

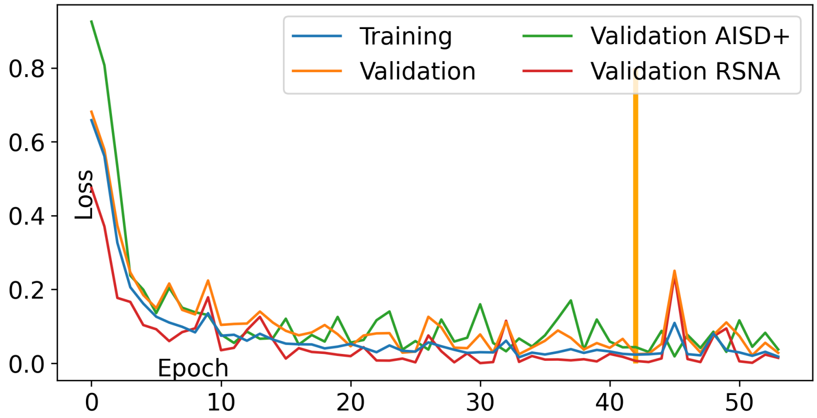


**(b)**

| | AUC ↑ | Accuracy ↑ | Precision ↑ | Recall ↑ | Specificity ↑ | F1-score ↑ |
|---|---|---|---|---|---|---|
| Chilamkurthy et al. (14) | $0.9422 \pm 0.010$ | $0.9425 \pm 0.010$ | $0.9205 \pm 0.010$ | $\mathbf{0.9463 \pm 0.010}$ | $0.9021 \pm 0.010$ | $0.9104 \pm 0.010$ |
| ResNet (15) | $0.9504 \pm 0.011$ | $0.9498 \pm 0.011$ | $0.9077 \pm 0.011$ | $0.9277 \pm 0.011$ | $0.8877 \pm 0.011$ | $0.9038 \pm 0.011$ |
| SE-ResNeXt (16) | $0.9557 \pm 0.012$ | $\mathbf{0.9530 \pm 0.012}$ | $0.9124 \pm 0.012$ | $0.8953 \pm 0.012$ | $0.8835 \pm 0.012$ | $0.9155 \pm 0.012$ |
| EfficientNet-B0 (17) | $0.9430 \pm 0.010$ | $0.9315 \pm 0.010$ | $0.9163 \pm 0.010$ | $0.9260 \pm 0.010$ | $0.8960 \pm 0.010$ | $0.9340 \pm 0.010$ |
| **Ours** | $\mathbf{0.9661 \pm 0.009}$ | $0.9521 \pm 0.009$ | $\mathbf{0.9379 \pm 0.009}$ | $0.9127 \pm 0.009$ | $\mathbf{0.9048 \pm 0.009}$ | $\mathbf{0.9376 \pm 0.009}$ |

**(c)**

| | AUC ↑ | Accuracy ↑ | Precision ↑ | Recall ↑ | Specificity ↑ | F1-score ↑ |
|---|---|---|---|---|---|---|
| Zunair et al. (18) | $0.8788 \pm 0.010$ | $0.9154 \pm 0.010$ | $0.8957 \pm 0.010$ | $0.9022 \pm 0.010$ | $0.8563 \pm 0.010$ | $0.8935 \pm 0.010$ |
| Solovyev et al. (19) | $0.9151 \pm 0.011$ | $0.9079 \pm 0.011$ | $0.9033 \pm 0.011$ | $0.8950 \pm 0.011$ | $0.8612 \pm 0.011$ | $0.8957 \pm 0.011$ |
| Pecoraro (20) | $0.8934 \pm 0.012$ | $0.9133 \pm 0.012$ | $0.8899 \pm 0.012$ | $0.8881 \pm 0.012$ | $0.8665 \pm 0.012$ | $0.9012 \pm 0.012$ |
| **Ours** | $\mathbf{0.9323 \pm 0.009}$ | $\mathbf{0.9320 \pm 0.009}$ | $\mathbf{0.9177 \pm 0.009}$ | $\mathbf{0.9312 \pm 0.009}$ | $\mathbf{0.8885 \pm 0.009}$ | $\mathbf{0.9189 \pm 0.009}$ |

**(d)**

**Fig. S5.** Experimental results for stroke subtype classification for comparison with state-of-the-art methods. (a) Dataset distributions for different categories. (b) Training and validation losses by epochs. (c) Quantitative testing results including AUC, accuracy, precision, recall, specificity, and F1-score on CQ500. (d) Quantitative testing results including AUC, accuracy, precision, recall, specificity, and F1-score on collected BGD-ISD.

### Supplementary: Ablation Study

**Table S3. Ablation study for segmentation module. (a) 2D U-Net vs 3D U-Net vs StrokeGNN. (b) Different choices on $k$ value, we choose 2 to 6 because model performance drops dramatically when $k = 6$. (c) Single-head vs multi-head (for ischemic and hemorrhagic, respectively).**

| | Dice ↑ | IOU ↑ | Precision ↑ | Recall ↑ | ASSD ↓ | Hausdorff ↓ |
|---|---|---|---|---|---|---|
| 2D U-Net (5) | 0.5557 | 0.3640 | 0.5883 | 0.5677 | 2.0 | 8.8 |
| 3D U-Net (5) | 0.5719 | 0.3765 | 0.6258 | 0.5822 | 2.0 | 8.4 |
| **StrokeGNN** | **0.6434** | **0.4507** | **0.7053** | **0.5983** | **1.8** | **7.8** |

**(a)**

| | Dice ↑ | IOU ↑ | Precision ↑ | Recall ↑ | ASSD ↓ | Hausdorff ↓ |
|---|---|---|---|---|---|---|
| $k = 2$ | 0.6403 | 0.4368 | 0.6944 | **0.6173** | **1.8** | 8.1 |
| $\mathbf{k = 3}$ | **0.6434** | **0.4507** | **0.7053** | 0.5983 | **1.8** | **7.8** |
| $k = 4$ | 0.6347 | 0.4349 | 0.6872 | 0.6044 | 2.1 | 8.3 |
| $k = 5$ | 0.6010 | 0.3976 | 0.6552 | 0.5777 | 2.4 | 8.9 |
| $k = 6$ | 0.5630 | 0.3710 | 0.6184 | 0.5671 | 2.6 | 9.1 |

**(b)**

| | Dice ↑ (ischemic) | Dice ↑ (hemorrhagic) |
|---|---|---|
| Single head | 0.5105 | 0.5971 |
| **Two heads** | **0.5793** | **0.6823** |

**(c)**

**Table S4. Ablation study for classification module. (a) 2D CNN vs 3D CNN. (b) No transfer learning (i.e., training from scratch) vs transfer learning.**

| | AUC ↑ | Accuracy ↑ | Precision ↑ | Recall ↑ | Specificity ↑ | F1-score ↑ |
|---|---|---|---|---|---|---|
| 2D CNN | 0.9412 | 0.9277 | 0.9058 | 0.9233 | **0.9244** | 0.9258 |
| **3D CNN** | **0.9522** | **0.9468** | **0.9176** | **0.9489** | 0.8866 | **0.9348** |

**(a)**

| | AUC ↑ | Accuracy ↑ | Precision ↑ | Recall ↑ | Specificity ↑ | F1-score ↑ |
|---|---|---|---|---|---|---|
| No transfer learning | 0.9250 | 0.8817 | 0.8632 | 0.9026 | 0.8627 | 0.9150 |
| **Transfer learning** | **0.9522** | **0.9468** | **0.9176** | **0.9489** | **8866** | **0.9348** |

**(b)**

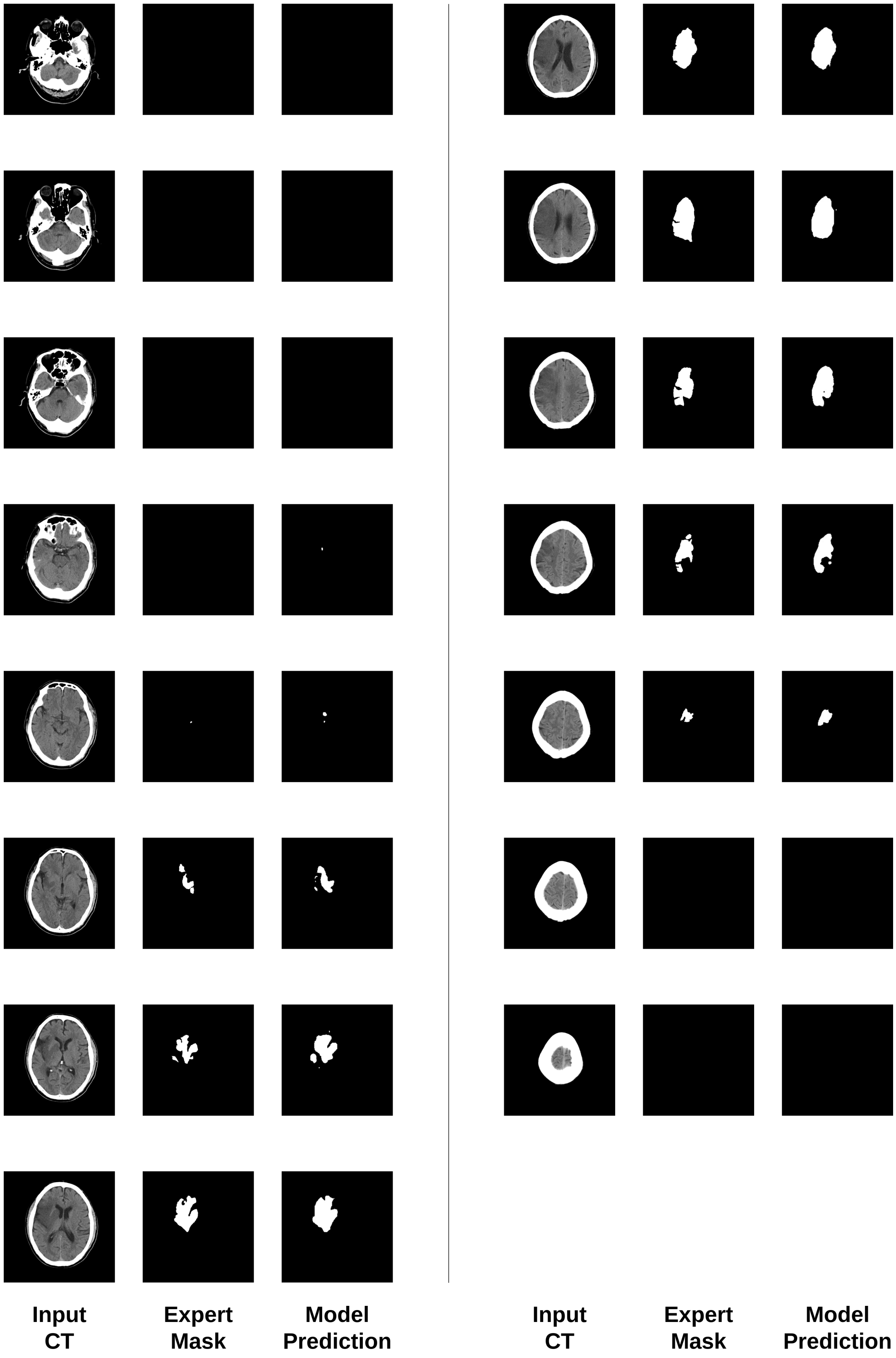


**Fig. S6.** Testing results for Sample 0072729 from the AISD dataset. The upper panel shows the segmentation output along with the input CT and expert mask. The predicted stroke type is classified as *ischemic stroke*, and the severity level is estimated as *severe*.

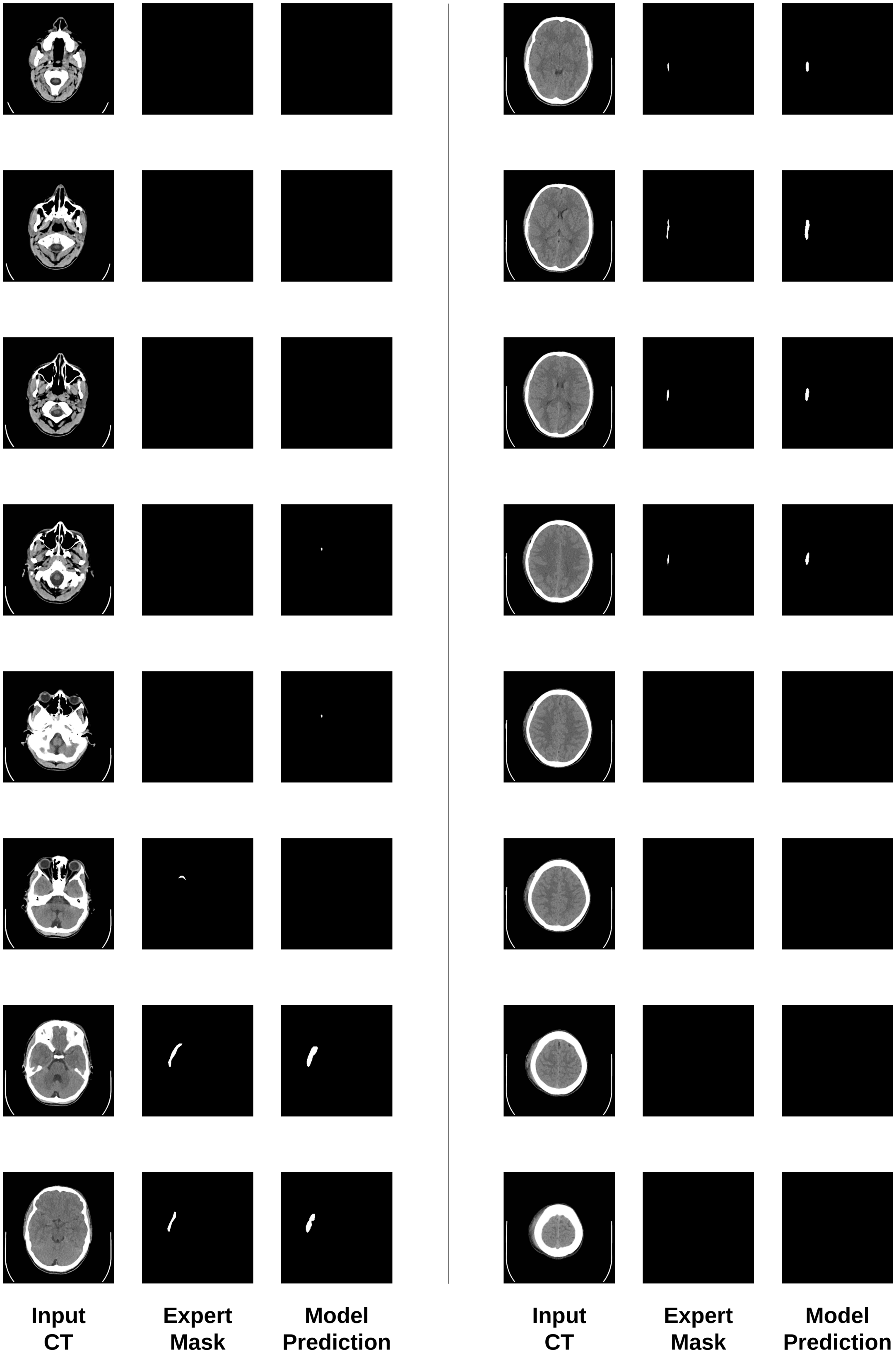


**Fig. S7.** Testing results for Sample 074 from the PhysioNet-ICH dataset. The upper panel shows the segmentation output along with the input CT and expert mask. The predicted stroke type is classified as *hemorrhagic stroke*, and the severity level is estimated as *minor*.

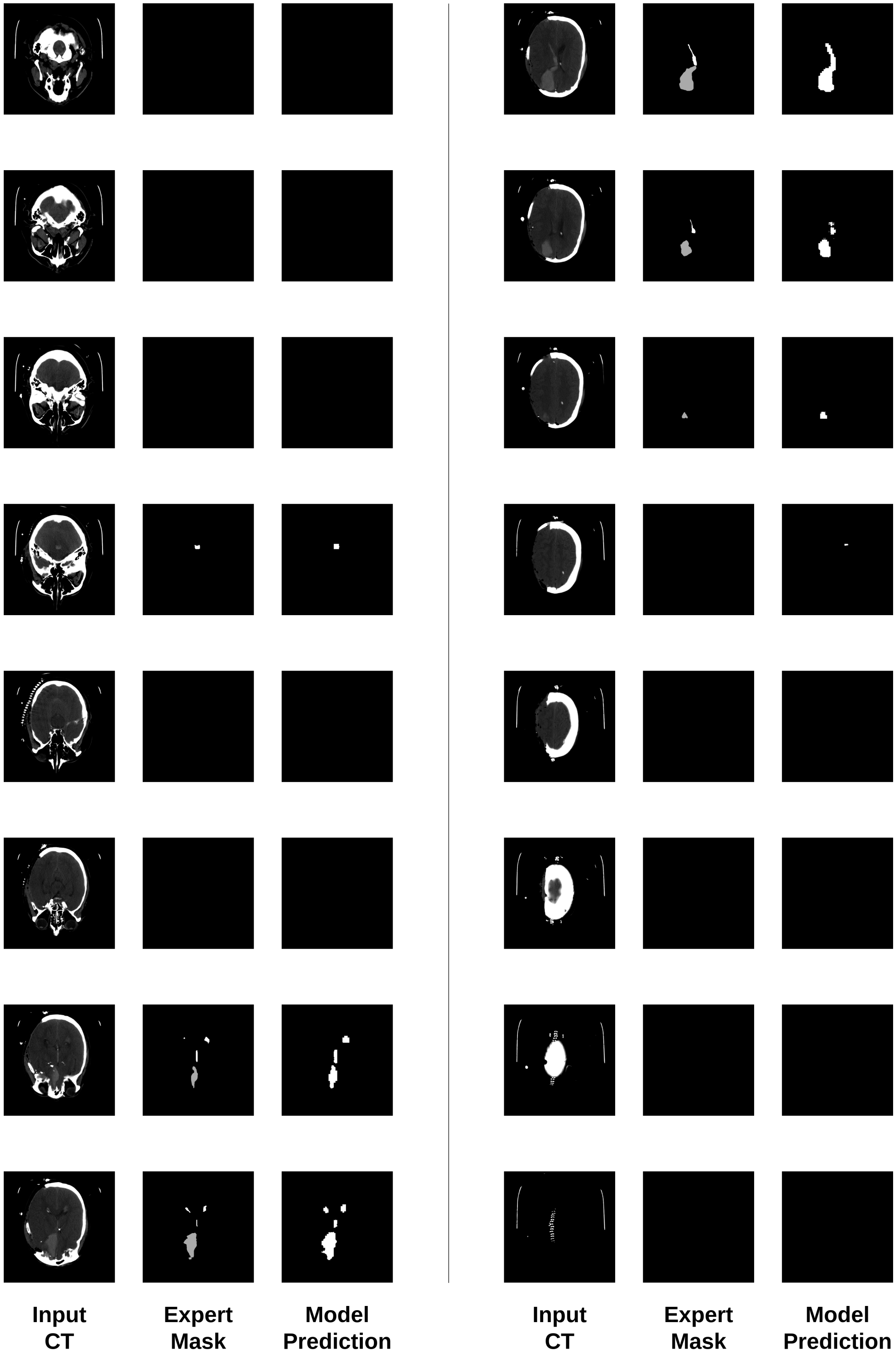


**Fig. S8.** Testing results for Sample 020 from the BHSD dataset. The upper panel shows the segmentation output along with the input CT and expert mask. The predicted stroke type is classified as *hemorrhagic stroke*, and the severity level is estimated as *moderate to severe*. Note that different colors in the expert mask column differentiate types of hemorrhages. In contrast, our model prediction provides a binary segmentation in order to maintain consistency with the labeling of other datasets.

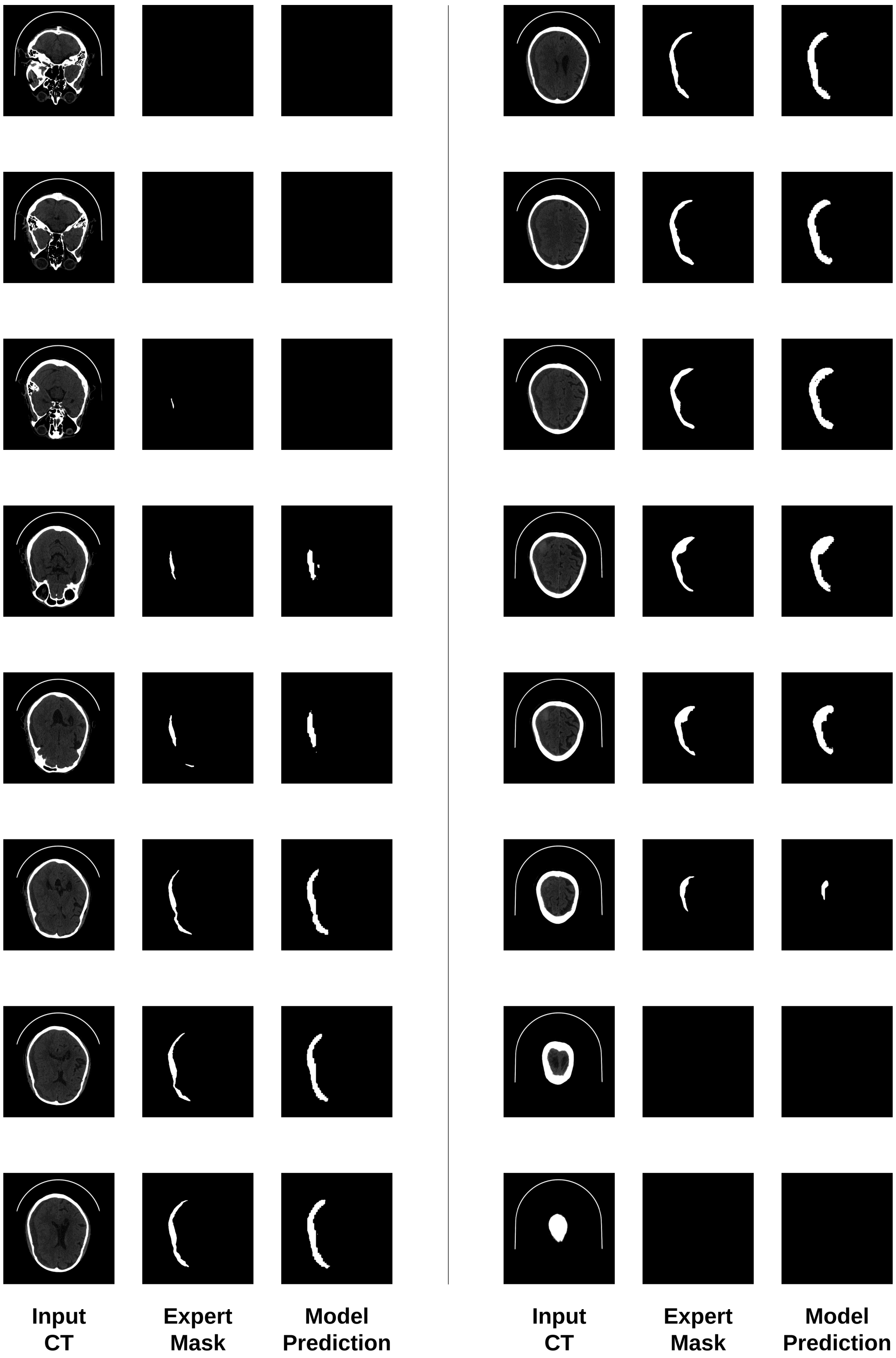


**Fig. S9.** Testing results for Sample 480 from the Seg-CQ500 dataset. The upper panel shows the segmentation output along with the input CT and expert mask. The predicted stroke type is classified as *hemorrhagic stroke*, and the severity level is estimated as *moderate*.

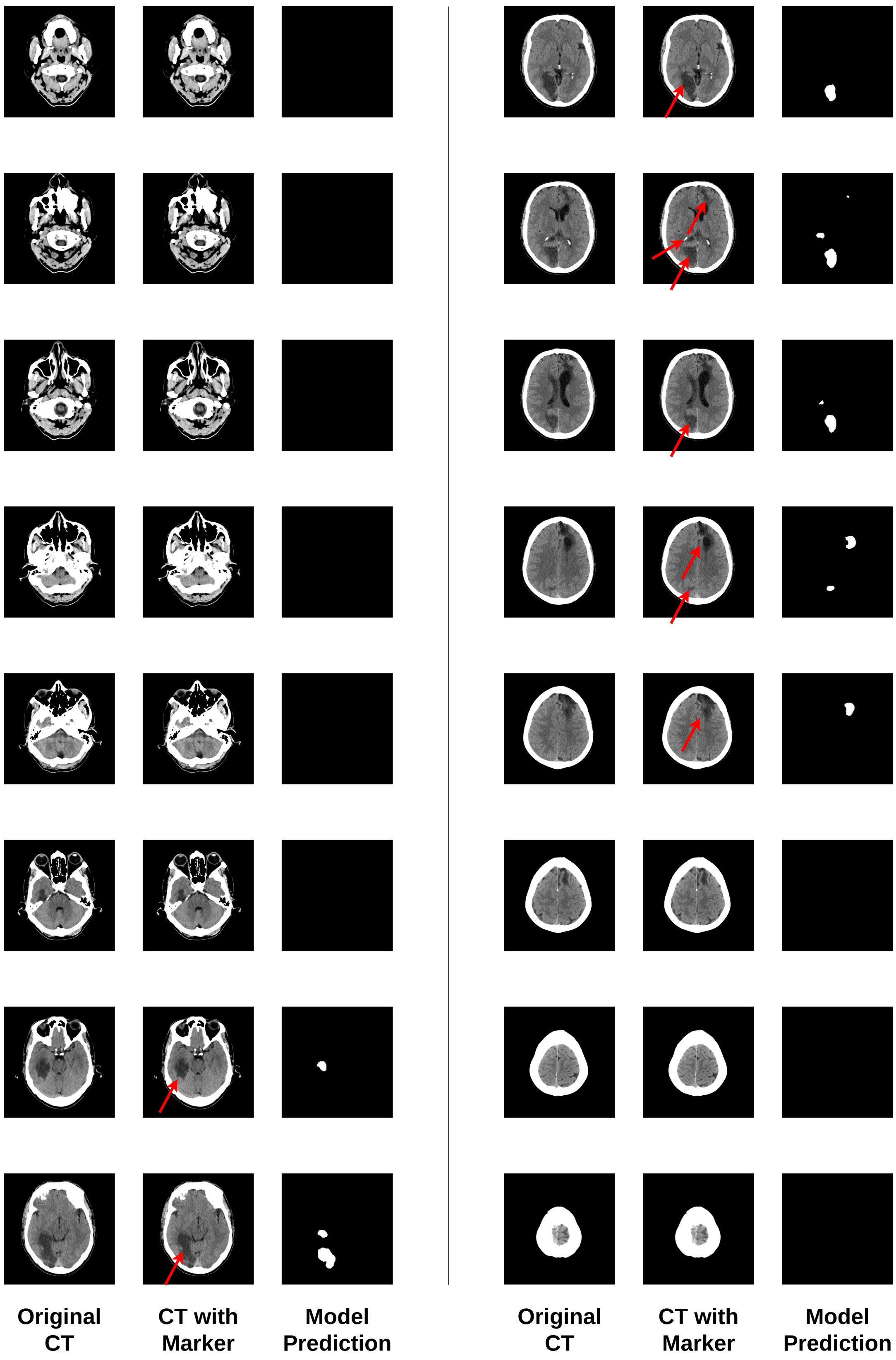


**Fig. S10.** Testing results for Sample 0001 from the collected BGD-ISDdataset. The upper panel shows the segmentation output along with the original CT and CT with marker indicating the potential lesion areas (note that the visual markers are not used for training or evaluation). The predicted stroke type is classified as *ischemic stroke*, and the severity level is estimated as *moderate*.

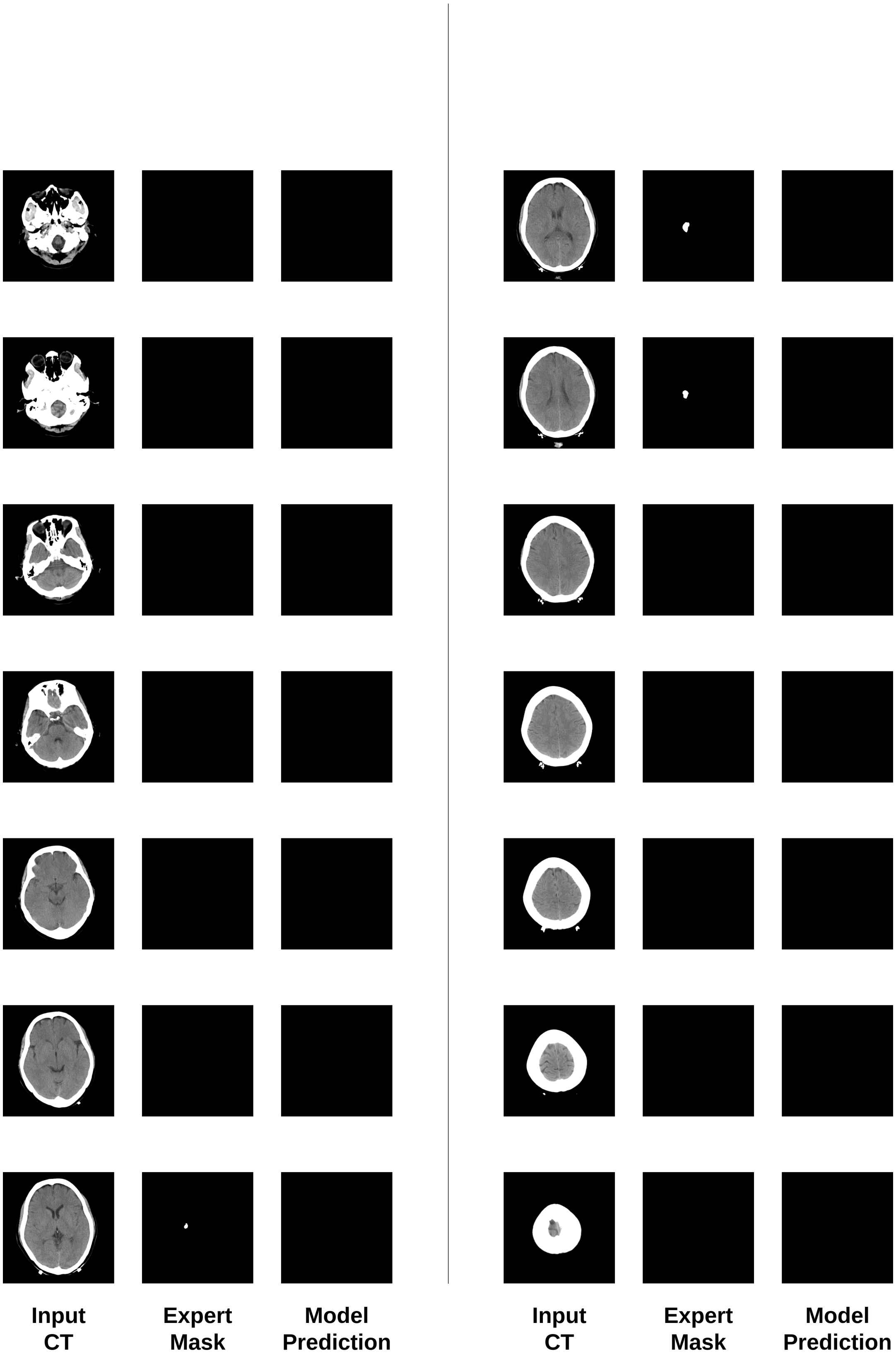


**Fig. S11.** Testing results for Sample 0073189 from the AISD dataset. The upper panel shows the segmentation output along with the input CT and expert mask. The predicted stroke type is classified as *no stroke*, and the severity level is estimated as *no stroke*.

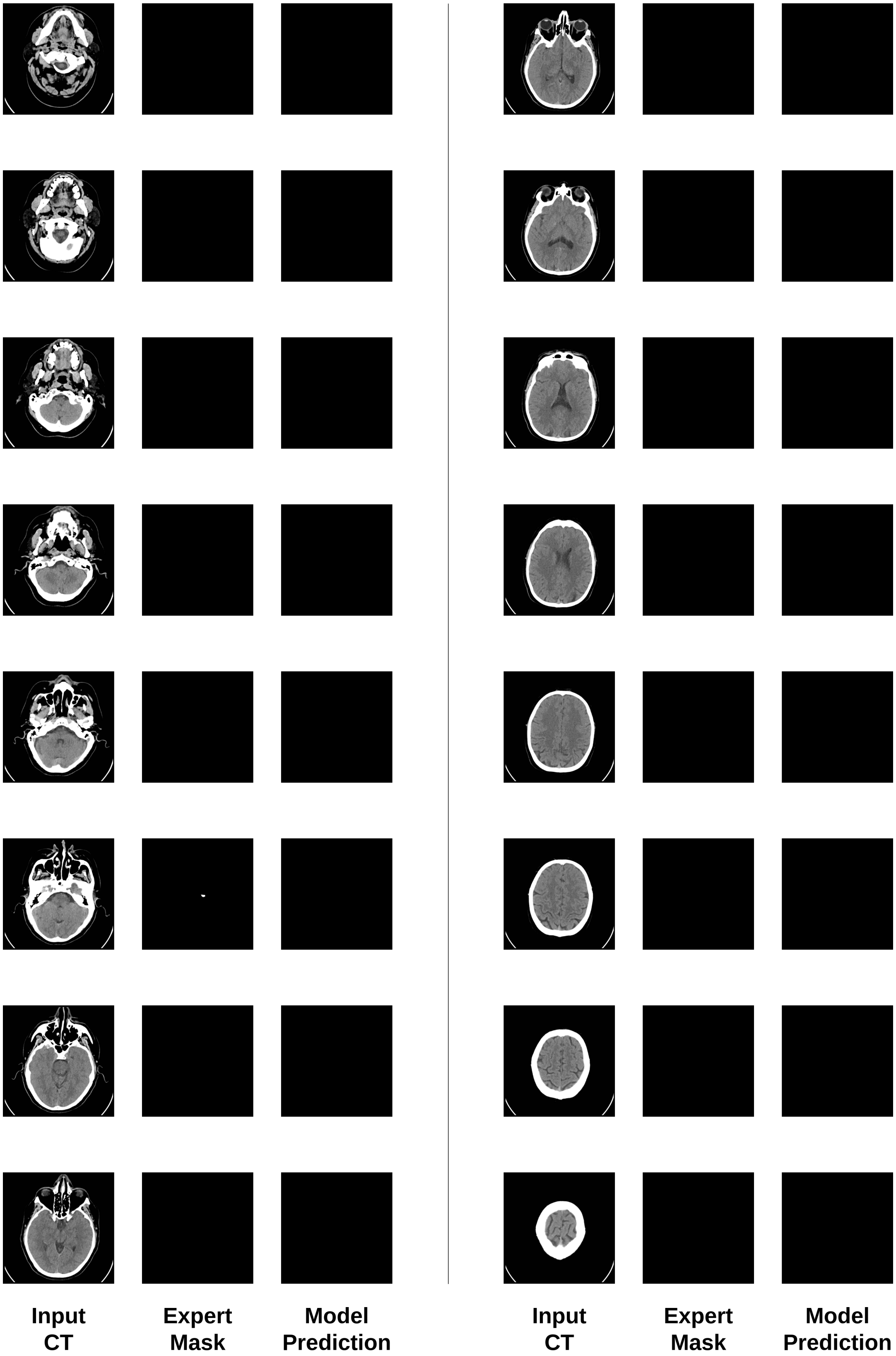


**Fig. S12.** Testing results for Sample 058 from the PhysioNet-ICH dataset. The upper panel shows the segmentation output along with the input CT and expert mask. The predicted stroke type is classified as *no stroke*, and the severity level is estimated as *no stroke*.

### Supplementary: Overview of Public Datasets

**Table S5. Brief overview of public datasets used in this article. More detailed overviews are shown on the next page.**

| Dataset Name | Download Link |
|---|---|
| AISD | https://github.com/GriffinLiang/AISD |
| PhysioNet-ICH | https://physionet.org/content/ct-ich/1.3.1/ |
| BHSD | https://huggingface.co/datasets/Wendy-Fly/BHSD |
| Seg-CQ500 | https://zenodo.org/records/8063221 |
| RSNA | https://www.kaggle.com/c/rsna-intracranial-hemorrhage-detection/data |
| CQ500 | http://15.206.3.216/dataset |

**Table S6. Detailed overviews of public datasets used in this article.**

| Acronym | Full Name | Download | Publication | Size | Description |
|---|---|---|---|---|---|
| AISD | Acute Ischemic Stroke Dataset | https://github.com/GriffinLiang/AISD | Symmetry-Enhanced Attention Network for Acute Ischemic Infarct Segmentation with Non-Contrast CT Images | 397 | The dataset includes 397 non-contrast CT (NCCT) scans from patients with acute ischemic stroke, all acquired within 24 hours of symptom onset. Each patient also received a diffusion-weighted MRI (DWI) within 24 hours post-CT. The NCCT scans (5mm slice thickness) are split into 345 for training/validation and 52 for testing. Ischemic lesions were manually annotated on NCCT using DWI as reference, with annotations reviewed by a senior clinician. |
| PhysioNet-ICH | PhysioNet-ICH | https://physionet.org/content/ct-ich/1.3.1/ | Intracranial Hemorrhage Segmentation Using a Deep Convolutional Model | 82 | The PhysioNet-ICH dataset includes 82 head CT scans from patients with traumatic brain injury (TBI), with slice-level annotations of intracranial hemorrhage (ICH) regions provided by two expert radiologists. Each slice is labeled for the presence of various hemorrhage subtypes and fractures. The dataset facilitates research in automated hemorrhage detection and clinical decision support. |
| BHSD | Brain Hemorrhage Segmentation Dataset | https://huggingface.co/datasets/Wendy-Fly/BHSD | A 3D Multi-Class Brain Hemorrhage Segmentation Dataset | 192 | The Brain Hemorrhage Segmentation Dataset (BHSD) consists of 192 CT volumes with pixel-level annotations and 1,980 unlabeled volumes, covering five types of intracranial hemorrhage (ICH). It supports 3D multi-class segmentation, enabling precise detection, localization, and quantification of ICH for clinical and research purposes. |
| Seg-CQ500 | Seg-CQ500 | https://zenodo.org/records/8063221 | Label-efficient deep semantic segmentation of intracranial hemorrhages in CT-scans | 51 | This dataset provides 3D segmentation masks of intracranial hemorrhages for 51 CT scans from the CQ500 dataset (http://headctstudy.qure.ai/dataset). Two trained radiologists from the Karolinska Institute in Stockholm annotated the scans to delineate hemorrhagic regions. |
| RSNA | RSNA Intracranial Hemorrhage Detection | https://www.kaggle.com/c/rsna-intracranial-hemorrhage-detection/data | Construction of a machine learn ing dataset through collaboration: the rsna 2019 brain ct hemorrhage challenge | 18938 | The RSNA Intracranial Hemorrhage Detection dataset includes over 25,000 head CT scans labeled for five subtypes of hemorrhage. Annotations were provided by expert radiologists, with each 2D slice labeled independently for the presence and type of hemorrhage. This large-scale dataset supports both classification and localization tasks in ICH diagnosis. |
| CQ500 | CQ500 | http://15.206.3.216/dataset | Development and Validation of Deep Learning Algorithms for Detection of Critical Findings in Head CT Scans | 491 | The CQ500 dataset comprises 491 head CT scans collected from multiple radiology centers in India, curated to support research in brain abnormality detection. Each scan includes expert annotations for a variety of critical findings, including intracranial hemorrhages, fractures, and mass effects. The dataset has been widely used to evaluate automated diagnostic models for emergency CT interpretation. |
